# Characterization of annual average traffic-related air pollution levels (particle number, black carbon, nitrogen dioxide, PM_2.5_, carbon dioxide) in the greater Seattle area from a year-long mobile monitoring campaign

**DOI:** 10.1101/2021.09.18.21263522

**Authors:** Magali N. Blanco, Amanda Gassett, Timothy Gould, Annie Doubleday, David L. Slager, Elena Austin, Edmund Seto, Timothy Larson, Julian Marshall, Lianne Sheppard

## Abstract

Growing evidence links traffic-related air pollution (TRAP) to adverse health effects. We designed an innovative and extensive mobile monitoring campaign to characterize TRAP exposure levels for the Adult Changes in Thought (ACT) study, a Seattle-based cohort. The campaign measured particle number concentration (PNC) to capture ultrafine particles (UFP), black carbon (BC), nitrogen dioxide (NO_2_), fine particulate matter (PM_2.5_), and carbon dioxide (CO_2_) at 309 stop sites representative of the cohort. We collected about 29 two-minute visit measures at each site during all seasons, days of the week, and most times of day during a one-year period. Validation showed good agreement between our BC, NO_2_, and PM_2.5_ measurements and regulatory monitoring sites (R^2^ = 0.68-0.73). Universal kriging–partial least squares models of annual average pollutant concentrations had cross-validated mean square error-based R^2^ (and root mean square error) values of 0.77 (1,177 pt/cm^3^) for PNC, 0.60 (102 ng/m^3^) for BC, 0.77 (1.3 ppb) for NO_2_, 0.70 (0.3 µg/m^3^) for PM_2.5_, and 0.50 (4.2 ppm) for CO_2_. Overall, we found that the design of this extensive campaign captured the spatial pollutant variations well and these were explained by sensible land use features, including those related to traffic.

**Synopsis:** We develop well-performing, long-term average pollutant exposure prediction models for epidemiologic application from an innovative and extensive short-term mobile monitoring campaign.

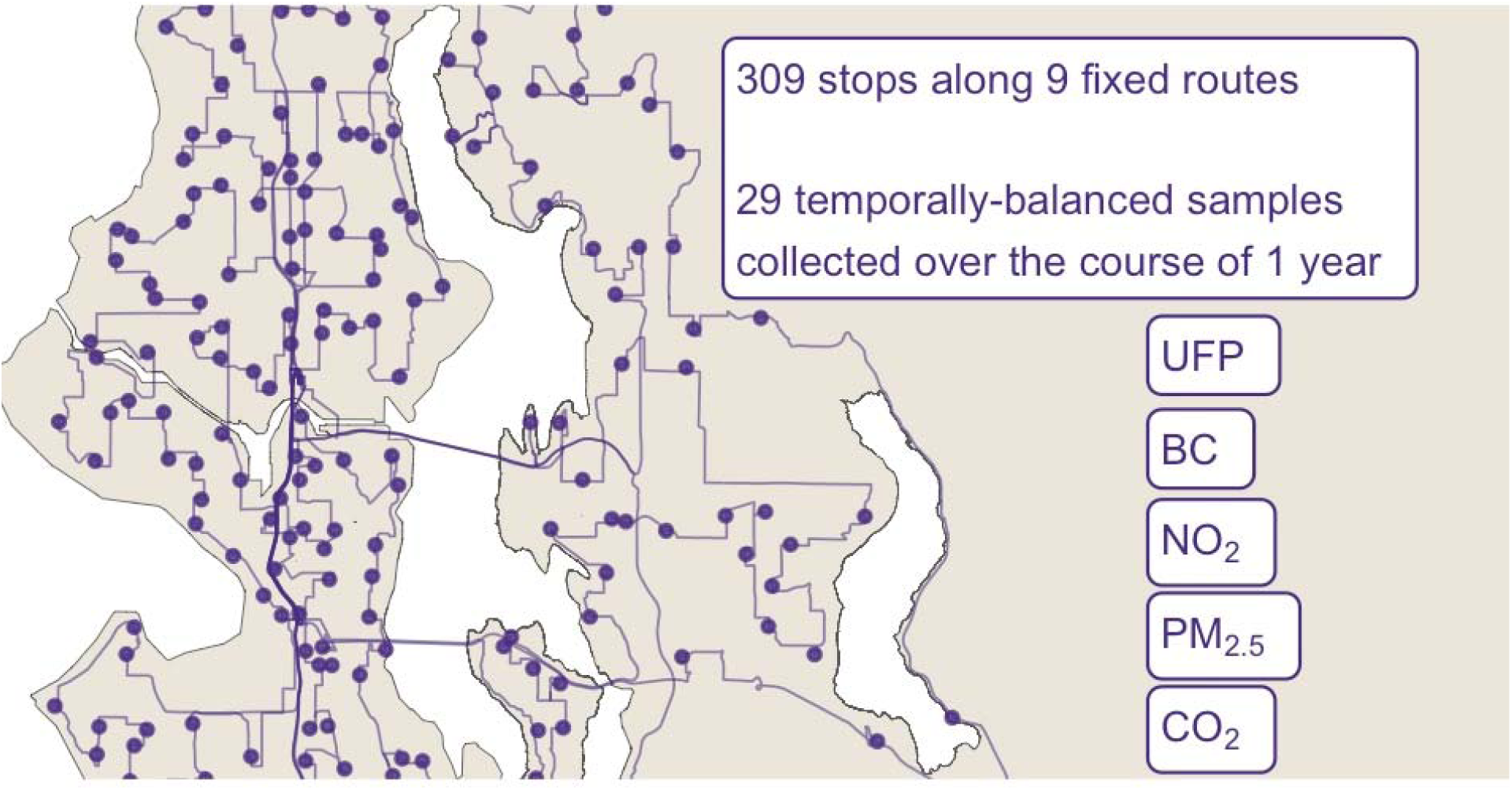

## 1 Introduction

An extensive body of evidence has linked air pollution to adverse health effects including respiratory, cardiovascular and mortality outcomes.^1^ Recent evidence has begun to link traffic-related air pollution (TRAP) exposure to cognitive function among various populations, including the elderly.^2–6^ While TRAP is a complex mixture that varies over time and space, pollutants include ultrafine particles (UFP; typically defined as aerodynamic diameter ≤ 100 nm), black carbon (BC), oxides of nitrogen including nitrogen dioxide (NO_2_), carbon dioxide (CO_2_), and carbon monoxide (CO).^7^ In particular, UFPs have increasingly been associated with important health outcomes including more neurotoxicity and systemic inflammation than larger particles.^8–14^

To date, however, much of the epidemiology air pollution research has been limited to the federally defined criteria air pollutants, monitored nationwide through the EPA’s regulatory Air Quality System (AQS) monitoring network. This network has monitored criteria pollutant levels throughout the US since the 1990s, and none specifically include UFPs.^15^ Furthermore, this network is spatially sparse and thus fails to capture the spatial variability of more quickly decaying pollutants, including many TRAPs.^16^ The Seattle Census Urbanized Area, for example, averages about 1 AQS monitor every 174 km^2^ (∼14 active monitors within a land area of about 2,440 km^2^), most of which measure fine particulate matter mass concentration with diameter of less than 2.5 µm (PM_2.5_) and BC.^17,18^

Mobile monitoring campaigns for assessing air pollution exposure have been used since at least the 1970s and have become increasingly common in recent years in an effort to address the limitations of traditional fixed site monitoring approaches.^19–25^ Typically, a vehicle is equipped with air monitors capable of measuring pollutants with high temporal resolution. Short-term sampling repeatedly occurs with this platform at predefined sites. Past work has shown that repeated short-term air pollution samples can be used to calculate unbiased long-term averages, thus reducing the need for continuous fixed-site monitoring.^19,20^ Because the sampling duration at individual sites can be quite short, campaigns can increase their spatial coverage with a single platform, thus making this approach more time- and cost-efficient than traditional fixed-site monitoring.

Still, the designs of past mobile monitoring campaigns have arguably limited their epidemiologic application. Importantly, most campaigns have sampled during limited time periods, for example, weekday business hours during one to three seasons.^21,26–28^ We previously showed that these limited sampling campaigns likely result in biased long-term human exposure estimates because they do not capture the high temporal variability of many TRAPs, and that the exact degree of bias varies (is not consistent) across site.^29^ Additionally, many campaigns have sampled along non-residential areas such as highways and industrial areas where air pollution levels may be much higher than the levels that most people are exposed to. Furthermore, most have collected non-stationary (mobile) on-road samples rather than stationary samples along the side of the road closer to participant residences. While non-stationary designs increase spatial coverage, further work is needed to demonstrate whether these are representative of residential human exposure levels.^21,30^ The additional bias that likely results from these limited sampling schemes is unclear.

To address the limitations of past campaigns, we designed an extensive, multi-pollutant mobile monitoring campaign to characterize TRAP exposure levels for the Adult Changes in Thought (ACT) study cohort. ACT is a long-standing, prospective cohort study that has been investigating aging and brain health in the greater Seattle area since 1995.^31^ The campaign measured TRAP at 309 stationary sites (stops) representative of the cohort in a temporally balanced approach throughout the course of a year. The goal of this paper is to describe the mobile monitoring design’s sampling methodology and TRAP measures collected, and to develop exposure predictions for later application to the ACT cohort. To the best of our knowledge, this is one of the most extensive mobile monitoring campaigns conducted in terms of the pollutants measured, the spatial coverage and resolution, and the campaign duration and sampling frequency.

## 2 Methods

Briefly, multiple pollutants including particle number concentration (PNC), BC, NO_2_, PM_2.5_, and CO_2_ were simultaneously measured with high quality instrumentation at 309 stop sites off the side of the road along fixed routes. Sites were representative of the cohort’s large spatial and geographical distribution throughout the greater Seattle area. A temporally balanced, year-long driving schedule that measured TRAP during all seasons, days of the week, and most times of the day enabled us to estimate unbiased annual average estimates at the site level. Details are described below.

### 2.1 Spatial Compatibility of the Selected Stop Sites and the ACT Cohort

We selected a mobile monitoring region in the greater Seattle, WA area that was roughly 1,200 land km^2^ (463 mi^2^; Figure 1). The monitoring region was composed of Census Tracts where the majority of the ACT cohort had historically resided between 1989-2018 (87% = 10,330/11,904 locations). This large region fell in western King County and southwest Snohomish County, and it included a variety of urban and rural areas with various land uses including residential, industrial, commercial, and downtown areas. We used the Location-Allocation tool in ArcMap (ArcGIS v. 10.5.1)^32^ to select 304 stops within the monitoring region that were representative of the ACT cohort (approximately one monitoring site per 33 participant locations; see Supplementary Information [SI] Note S1 for details). Stops were spatially distributed so that they would cover all parts of the monitoring region. The exact sites selected were meant to minimize the distance between the monitoring and cohort locations. Five additional stops were collocations at nearby regulatory air quality monitoring sites measuring pollutants similar to our platform (see below). In total, there were 309 stops. The average (SD) distance between a cohort location and the nearest monitoring stop was 611 (397) m. The monitoring stops and cohort locations had similar distributions of various TRAP-related covariates (e.g., proximity to roadways, airport, railyard), indicating good spatial compatibility (SI Figure S1).^33^

**Figure 1.**
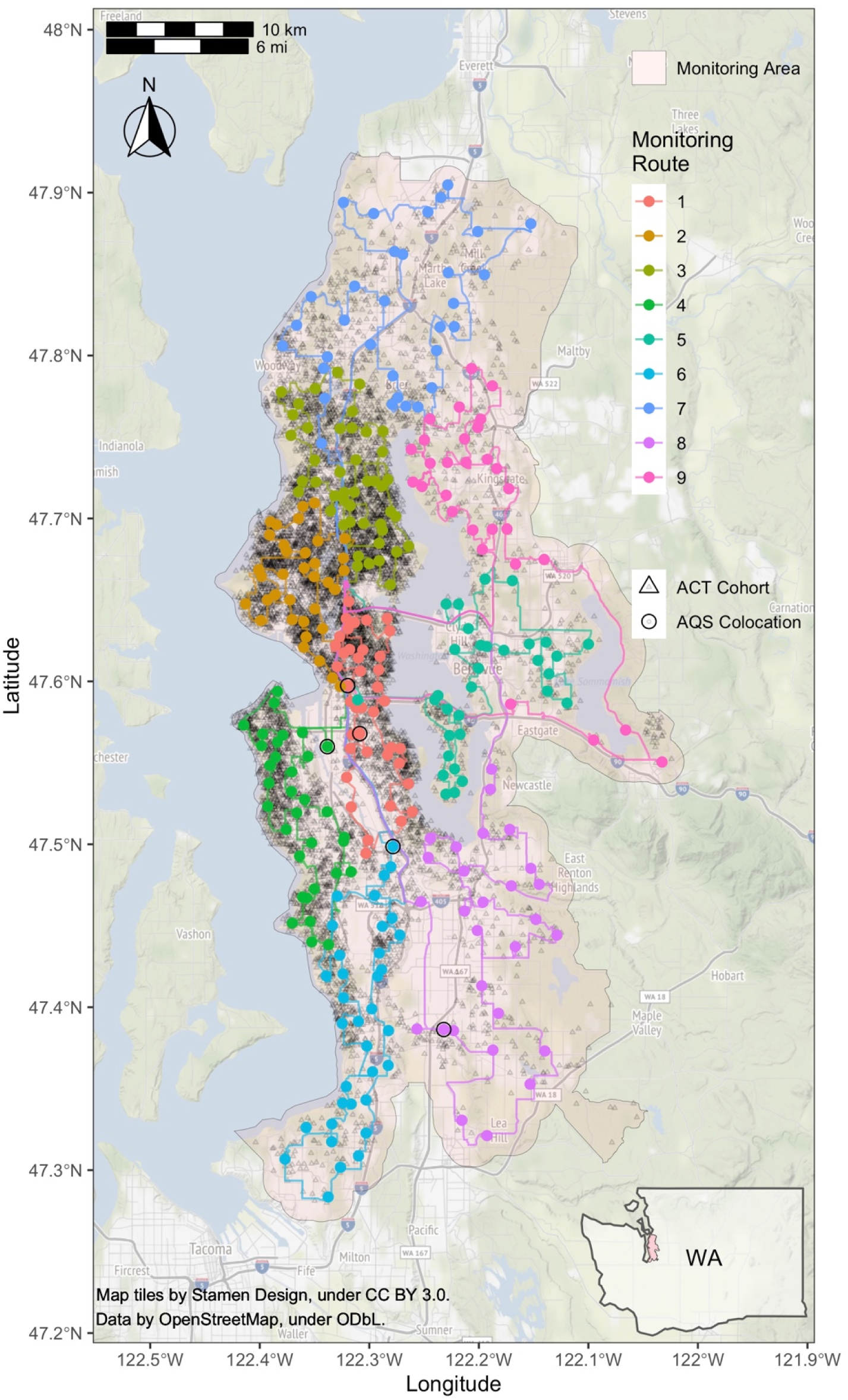
Mobile monitoring routes (n=309 stops along 9 routes) and jittered ACT cohort locations (n=10,330 unique locations). Inset map shows the monitoring area within Washington (WA) state.

### 2.2 Fixed Routes

We used ArcMap’s Network Analyst New Route tool^32^ and Google Maps^34^ to develop nine fixed routes based on the 309 stop monitoring sites. Each route ranged from 75-168 km (47-104 miles) in length and had 28-40 stops (SI Table S1). All routes started and ended at the University of Washington and were intended to maximize residential driving coverage (i.e., reduce highway driving and driving on the same roads). Routes were downloaded from Google Maps to a smart phone and Garmin GPS Navigation System, and navigation was set to replicate the same route each time regardless of traffic conditions.

### 2.3 Sampling Schedule

Sampling was conducted from March 2019 through March 2020 during all seasons and days of the week between the hours of 4 AM and 11 PM. Our previous work has shown that this balanced but slightly reduced sampling schedule taking driver safety and operational logistics into consideration should still generally produce unbiased annual averages.^29^ This work further showed that the temporal sampling design rather than the visit sampling duration has the largest impact on the accuracy of the annual average estimates, and that common sampling designs like weekday business and rush hours regularly produce more biased annual averages. To increase temporal coverage, routes were started at different times of the day and driven in both clockwise and counterclockwise directions. A single route was driven each day (∼4-8 drive hours). Make-up site visits were conducted throughout the study to resample sites with missing readings (i.e., due to instrumentation or driver errors). Make-up visits occurred during similar times as the originally scheduled sampling time (i.e., season, day of the week, general time of day). Twenty-eight two-minute samples were scheduled to be collected at each stop site while the vehicle was parked along the side of the road. This design choice was justified by our additional analyses of one-minute data from a near-road and a background regulatory site in Seattle. These analyses showed that at least 25 two-minute samples were sufficient to produce annual average estimates with a low average percent error (See SI Figure S2). Furthermore, there was only a negligible improvement in annual average estimates when the sampling duration was extended from 2 to 60 minutes.

### 2.4 Data Collection

We equipped a Toyota Prius hybrid vehicle with fast-response (1-60 sec), high-quality instrumentation that measured various particle and gas pollutants. Pollutants included BC (AethLabs MA200), NO_2_ (Aerodyne Research Inc. CAPS), PM_2.5_ (Radiance Research M903 nephelometer), CO_2_ (Li-Cor LI-850), and PNC with various instruments, including two TSI P-TRAK 8525’s (one unscreened – the primary instrument in this analysis, and one with a diffusion screen), a TSI NanoScan 3910, and Testo DiSCmini. PNC serves as a surrogate for UFP since most particles by count are smaller than 100 nm.^35^ CO measurements were also collected, but these were not included in this analysis because they did not meet our quality standards. The platform additionally collected temperature, relative humidity, and global positioning with real-time tracking. See SI Table S2 for instrumentation details, including the manufacturer-reported size ranges for the four PNC instruments. We had duplicates (back-ups) of every instrument type that were periodically collocated for quality assurance purposes (see Quality Assurance and Quality Control). SI Note S2 and Figures S3-S4 have additional details on the platform configuration and data collection procedures.

### 2.5 Quality Assurance and Quality Control

We conducted various quality assurance and quality control (QAQC) activities throughout the study period to ensure the reliability and integrity of our data. Activities included calibrating gas instruments; checking particle instruments for zero concentration responses; assessing collocated instruments for agreement; inspecting time series data for concentration pattern anomalies; and dropping readings associated with instrument error codes or those outside the instrument measurement range. SI Section S1.3 has additional details.

### 2.6 Site Visit Summaries

All data analyses were conducted in R (v 3.6.2, using RStudio v 1.2.5033; see SI Note S3 for computing details).^36^

We calculated the median pollutant concentrations for each two-minute site visit. While means can be highly influenced by large concentration deviations (which may be important in some settings), medians are more robust to outliers and may better capture the typical values of skewed data.

We estimated PM_2.5_ concentrations from nephelometer readings using a calibration curve fit to regulatory monitoring data between 1998-2017 (SI Equation S1). Nephelometer light scattering is strongly correlated with PM_2.5_ and has been used in the Puget Sound region to monitor air quality since 1967.^37^ We fit the model using daily average measurements from nine non-industrial regulatory air monitoring sites in the region where both PM_2.5_ (using federal reference methods) and nephelometer light scattering data were collected. We excluded the years 2008-2009 due to nephelometer instrumentation issues noted by the local regulatory agency. The model’s leave-one-site-out cross-validated R^2^ and root mean square error (RMSE) were 0.92 and 1.97 µg/m^3^, respectively.

Site visit medians and annual averages for BC, NO_2_ and PM_2.5_ estimated from these data were compared against estimates from the five regulatory air monitoring collocation sites.

### 2.7 Spatial and Temporal Variability

We ran analysis of variance (ANOVA) models for each pollutant to characterize the relative variability of the site visit level data over space, time, and within site. The independent variables for each pollutant model were the site (n=309), season (n=4), day of the week (n=7), and hour of the day (n=21), while the dependent variable was median visit concentrations.

### 2.8 Estimation of Annual Averages

We calculated winsorized annual average concentrations for each site such that concentrations below the 5^th^ and above the 95^th^ quantile concentration were substituted with the 5^th^ and 95^th^ quantile concentration, respectively (mean of winsorized medians). This was done to reduce the influence of large outlier concentrations on the annual average. In sensitivity analyses, we calculated non-winsorized averages (mean of medians) and medians (median of medians).

### 2.9 Annual Average Prediction Models

Development of annual average prediction models allows the predictions to be used for epidemiologic inference. The data were randomly split into a training-validation (90%, n=278 sites) and a test (10%, n=31 sites) set. The training-validation set was used to select the 191 geographic covariate predictors (e.g., land use, roadway proximity) that had sufficient variability and a limited number of outliers from 350 original covariates (see SI Notes S5 for details). These were summarized using pollutant-specific partial least squares (PLS) regression components. We built pollutant-specific universal kriging (UK) models for annual average concentrations, using log-transformed concentrations as the dependent variable and the first three geocovariate PLS principal components as the independent variables (Equation 1). We used UK rather than land use regression (LUR) alone since UK uses geospatial smoothing to capture any residual spatial correlation not otherwise captured by LUR. We selected the kriging variogram model for the geostatistical structure using the `fit.variogram` function in the `gstat`^38^ R (v 3.6.2, using RStudio v 1.2.5033)^36^ package.

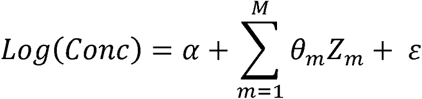

*Equation 1. Universal kriging with partial least squares models for annual average pollutant concentrations. Conc is the pollutant concentration, Z*_*m*_ *are the PLS principal component scores (M=3), α and θ*_*m*_ *are estimated model coefficients, and ε is the residual term with mean zero and a modeled geostatistical structure*.

We used RMSE and mean square error (MSE) -based R^2^ to evaluate the performance of each pollutant model on the native scale using ten-fold cross-validation and test sites. We used MSE-based R^2^ instead of traditional, regression-based R^2^ because it evaluates whether predictions and observations are the same (around the one-to-one line) such that it assesses both bias and variation around the one-to-one line. Regression-based R^2^, on the other hand, solely assesses whether pairs of observations are linearly associated, regardless of whether observations are the same or not.

## 3 Results

### 3.1 Data Collected

After dropping stop concentrations that did not meet the quality assurance standards (0.61%), the final analyses included over 70,000 two-minute median stop samples (almost 9,000 samples per instrument) collected over the course of 288 drive days from 309 monitoring sites (Table S7). Sites were sampled an average of 29 times, ranging from 26 to 35 times. Due to the logistical constraints of sampling 309 sites with one platform along nine fixed routes, some sites were visited fewer times of the day than other sites, though sampling times were still well distributed throughout the day (e.g., morning [e.g., 7 AM], afternoon [e.g., 3 PM] and evening [e.g., 8 PM]; see SI Figure S7). SI section S2.1 Site Visits has additional details on the visit-level pollutant concentrations used to estimate site annual averages.

### 3.2 Collocations at Regulatory Monitoring Sites

Median two-minute BC, NO_2_ and PM_2.5_ measurements from mobile monitoring stops were generally in agreement with measurements from regulatory sites (MSE-based R^2^: BC = 0.69, NO_2_ = 0.71, PM_2.5_ = 0.61; SI Figure S12). Annual average estimates from our mobile monitoring campaign measurements were similar to annual average estimates from comparable two-minute samples at regulatory monitoring sites used as collocations, and these were in moderate agreement with true annual average concentrations at those sites (based on all of the available data during the study period; SI Figure S13).

### 3.3 Spatial and Temporal Variability

Pollutant-specific ANOVA models of winsorized site visit concentrations indicated most of the concentration variability occurred within sites, rather than across sites or over time (SI Figure S14). After accounting for time and site, PNC from the P-TRAK instrument had the highest within-site variability (82% of the total), followed by PM_2.5_ (87%), BC (80%), CO_2_ (70%), and lastly, NO_2_ (66%). CO_2_ (27%) had the most temporal variability, followed by NO_2_ (24%), BC (16%), PM_2.5_ (13%), and PNC (6%), respectively. Finally, PNC (12%) had the most spatial variability, followed by NO_2_ (10%), BC (4%), CO_2_ (3%) and PM_2.5_ (<1%), respectively. Unlike other pollutants, PNC had more spatial than temporal variability. SI Figure S14 shows similar results for other PNC instruments.

### 3.4 Annual Average Estimates

Estimated annual average pollutant concentrations across all monitoring sites are shown in SI Figure S15. There was a 5- to 6-fold difference between the lowest and highest site concentrations of PNC, NO_2_, and BC. On the other hand, PM_2.5_ had a 2-fold difference across sites, while CO_2_ varied little across sites. Among PNC instruments, the screened P-TRAK measured the lowest concentrations and had the smallest variability; the P-TRAK, which did not screen out particles below 36 nm, had the second-highest averages with approximately double the values and more variability. The DisSCmini and Nanoscan had higher medians, more variability, and more outlying annual average concentrations. SI Figures S16-S17 map these concentrations. The locations with the highest BC, NO_2_, and PNC concentrations were near the Seattle urban core. High PNC concentration sites were additionally located at more southern locations near the area’s major airport, the Seattle-Tacoma (Sea-Tac) International Airport. Sites with elevated PM_2.5_ and CO_2_ levels were dispersed throughout the monitoring region.

### 3.5 Prediction Models

Based on the training-validation set, the first three PLS principal components captured between 49-51% of the observed concentration variability for each pollutant model. Loadings from the first PLS principal component indicated that normalized difference vegetation index (NDVI), length of bus routes, major roadways, land development, population density, and truck routes were strong predictors of air pollution in the region, with some pollutants, for example PNC, being more influenced by these land features (SI Figure S18). Cross-validated MSE-based R^2^ (and RMSE) values for UK-PLS models were 0.77 (1,177 pt/cm^3^) for PNC, 0.60 (102 ng/m^3^) for BC, 0.77 (1.3 ppb) for NO_2_, 0.70 (0.3 µg/m^3^) for PM_2.5_, and 0.51 (4.2 ppm) for CO_2_ (SI Table S9). In the independent test set, these results differed somewhat with estimates of MSE-based R^2^ (and RMSE) of 0.78 (815 pt/cm^3^) for PNC, 0.80 (60 ng/m^3^) for BC, 0.84 (0.9 ppb) for NO_2_, 0.73 (0.3 µg/m^3^) for PM_2.5_, and 0.77 (2.7 ppm) for CO_2_. Sensitivity analyses using mean of medians and median of medians annual averages performed similar or slightly lower due to changes in the number of influential points and/or reduced overall variability (SI Table S9). These model performances are reflected in the generally good agreement between the estimates and cross-validated predictions (Figure S19). All PNC instruments do show a few underpredicted observations.

Model predictions for the monitoring region are shown in Figure 2 (predictions from additional PNC instruments are shown in Figure S20). While PM_2.5_ and CO_2_ are fairly spatially homogeneous, PNC, BC, and NO_2_ (traditional TRAPs) show higher concentrations in the urban core and along major roads. In addition, PNC shows higher concentration near the area’s major airport. All the PNC instruments reflect this broad pattern, although there are differences across instruments in the areas with the highest predicted concentrations.

**Figure 2.**
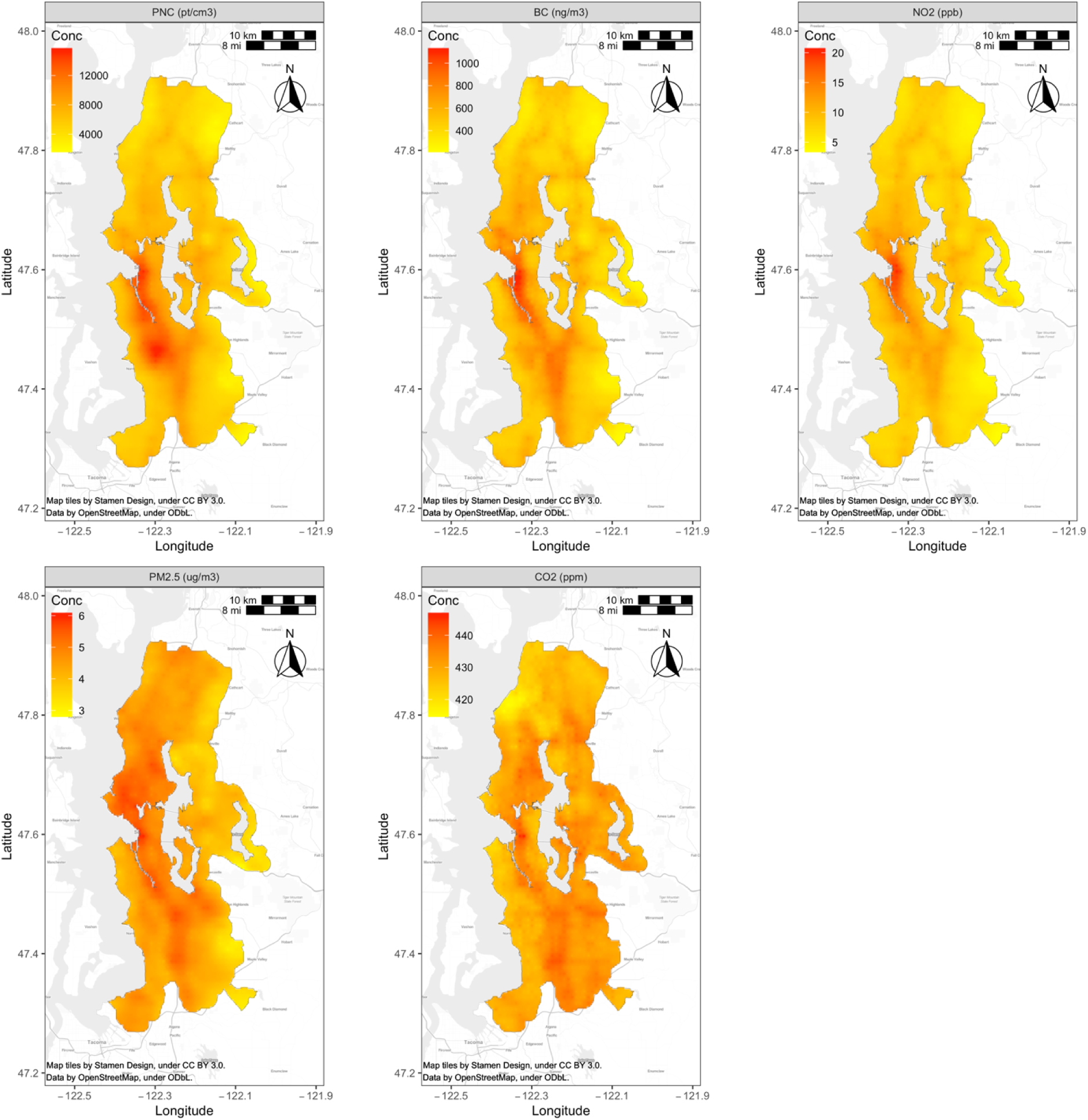
UK-PLS pollutant predictions for the monitoring region.

Pearson correlation coefficients (R) for pollutant model predictions at the 309 monitoring sites and all instruments are shown in SI Figure S21. Different PNC instruments were generally well correlated with each other (R = 0.85-0.97). Overall, PNC from the P-TRAK, BC, and NO_2_ were well correlated with each other (R = 0.81-0.92), and moderately correlated with PM_2.5_ and CO_2_ (R = 0.39-0.70). CO_2_ and PM_2.5_ were moderately correlated with each other (R = 0.46). The biggest deviations from a linear association were evident for the predicted high concentrations from the DiSCmini; this was particularly apparent in its relationship with BC, NO_2_, PM_2.5_, and CO_2_.

## 4 Discussion

In this paper, we describe the design of an innovative mobile monitoring campaign specifically developed to estimate unbiased, highly spatially resolved, long-term TRAP exposures in an epidemiologic cohort. To date, this is one of the most extensive mobile monitoring campaigns conducted in terms of the pollutants measured (five pollutants measured with eight different instruments, not including CO) spatial coverage (∼1,200 land km^2^), sampling density (309 monitoring sites along 9 routes, or 1 monitor every 3.9 land km^2^), and sampling frequency (7 days a week; 288 days over a one-year period) and duration (∼5 driving hours per day between the hours of 4 AM – 11 PM). The spatial resolution achieved by this campaign was significantly greater than would be expected from fixed regulatory monitoring approaches. We had one monitor per 3.9 km^2^ of land area rather than 183 km^2^ (6 regulatory sites in the monitoring area), almost a 50-fold increase. The average (SD) distance from an ACT cohort location to the nearest monitoring site was 611 (397) m rather than 5,805 (2,805) m to an AQS site, almost a ten-fold difference. Monitor proximity to prediction (i.e., cohort) locations, both in terms of geographic and covariate distance, is an important determinant of accurate exposure assessment.^39,40^ Additionally, we previously showed that the extensive temporal sampling of this campaign across hours, days of the week and seasons is expected to produce more accurate and unbiased annual average estimates as compared to more common campaigns with reduced sampling.^29^

A unique aspect of this campaign was the collection of stationary samples along the side of the road. While most other campaigns have only collected non-stationary, on-road samples, various studies have shown that mobile samples are generally higher in concentration than stationary samples.^21,41–44^ The completion of our stationary and non-stationary campaign positions us to conduct future work on how non-stationary data may be used responsibly for epidemiologic applications. Among the relatively few campaigns that have collected stationary rather than mobile samples alone, most have sampled for longer than two minutes (about 15-60 minutes per stop).^45^ Our analyses indicated that shorter sampling periods produce comparably good estimates without adding excessive amounts of stationary sampling time to mobile monitoring campaigns (See SI Figure S2). Our use of a hybrid vehicle meant that the vehicle’s engine was off and it operated by battery during stop sampling periods, thus reducing the possibility of self-contamination.

ANOVA model results indicate differences across pollutants in terms of their spatial and temporal variability. This finding is particularly relevant for short-term mobile monitoring campaigns, which could design their campaigns to adequately capture the variability of the pollutants of interest. These findings suggest that repeated sampling at any given site is crucial since most of the variability for all measured pollutants was seen within sites, even after adjusting for time. Following that, all pollutants other than PNC had relatively more temporal than spatial variability. Campaigns measuring these pollutants may thus benefit by inclusion of more temporally-balanced site visits. PNC, on the other hand, has slightly more spatial than temporal variability suggesting that both are important. The implementation of these concepts for epidemiologic exposure assessment should translate to reduced exposure misclassification. Overall, our results are in line with past literature that has shown differing spatial and temporal contrasts across pollutants,^46,47^ though our work increases the robustness of these findings using a more spatially resolved, multi-pollutant dataset that includes less commonly measured PNC.

The findings from this campaign demonstrate the region’s generally low air pollution levels. The range of annual concentrations across sites for PM_2.5_ (3.4-7.2 µg/m^3^) and NO_2_ (3.9-23 ppb) were well below the National Ambient Air Quality Standards (NAAQS) annual average levels of 12 µg/m^3^ and 53 ppb, respectively.^48^ Annual PNC (∼7,000 pt/cm^3^) and BC (∼600 ng/m^3^) site concentrations were lower than what others have reported in cities throughout the world where mean study values range from roughly 6,000-64,000 PNC pt/cm^3^ and 400-14,000 BC ng/m^3^ (PNC^21,42,43,49–63^; BC^19,21,43,52,53,58,63–75^). While CO_2_ site concentrations (417-455 ppm) were above the 2019 global average of 412 ppb,^76^ they were in line with past work noting elevated carbon footprint levels in dense, high-income cities and affluent suburbs.^77,78^ Still, the high concentration variability seen across sites for pollutants like PNC, BC and NO_2_ suggests that future epidemiological analyses may have more power to observe health effects from these pollutants than those that are less spatially variable, for example PM_2.5_ and CO_2_.

The similarity between BC, NO_2_ and PM_2.5_ measurements from our campaign and collocated regulatory monitoring sites confirms that our campaign estimates were generally accurate. Some of the discrepancies between the two monitoring approaches may be due to differences in the sampling instrumentation, the exact sampling location, and quality assurance and quality control procedures. While we were unable to compare CO_2_ or PNC measurements to regulatory observations, duplicate instrument collocations generally showed good agreement (SI Figure S6). Additionally, CO_2_ instruments were regularly calibrated and PNC instruments completed zero checks (SI Table S3, Figure S5).

We observed elevated annual average pollutant levels near areas with low green space (as quantified by normalized difference vegetation index [NDVI]), bus routes, major roadways, and impervious surfaces. These findings are generally in line with past work.^79^

While future mobile monitoring campaigns may be guided by the design and findings from this study, it’s notable that the unique geographical, meteorological and source characteristics of different airsheds may produce slightly different results. These results do highlight, however, the importance of collecting multi-pollutant measurements, particularly in urban or other areas characterized by major emission sources such as airports or railroad systems, which may be important contributors to local and/or regional air pollution levels. This is particularly true for PNC given the limited monitoring data available and its unique spatial and temporal patterns. More generally, multi-pollutant exposure assessment is a growing interest in the field of air pollution epidemiology,^46,80–83^ and something that we are positioned to make a meaningful contribution to in future work.

While UFPs are generally characterized as particles under 100 nm in diameter, this definition is not standardized and varies from instrument to instrument as well as study to study. Since most particles by count are in the smaller size range with few above 100 nm,^35^ PNC should adequately characterize UFPs. Moreover, the collection of PNC from multiple instruments in a field setting is unique to this study. PNC measures from different instruments were strongly correlated with each other, and they produced broadly similar spatial surfaces, strengthening our confidence in the quality of our measurements. Differences in the reported PNC levels across instruments, however, can be attributed to multiple factors including differences in technology, each technology’s unique particle size detection efficacy, and built-in calibration (if present), all of which impact the reported particle size ranges and concentrations of each instrument. Differences across PNC instruments in the predicted absolute concentrations as well as overall spatial surfaces highlight these differences. By comparing PNC levels from the unscreened and screened P-TRAK, for example, we see that roughly half of the measured (and predicted) particles are between 20-36 nm (SI Figure S20). Furthermore, these smaller particles are more concentrated near the area’s major airport, the Sea-Tac International Airport. The DiSCmini also captures this rise in PNC near the airport but shows much lower relative concentrations elsewhere, suggesting it measures smaller particles well. Reasons could include the different measurement technology as well as the manufacturer’s reported lower particle size cut of 10 nm. The NanoScan total concentration, on the other hand, reports concentrations that are roughly 50% higher than the unscreened P-TRAK, with elevated PNC levels near the airport, but also in other parts of the monitoring region, including south of the airport along major roadways and at the Seattle urban core. Elevated PNC levels are thus predicted from the NanoScan in a larger area of the monitoring region.

It is an open question whether the use of different PNC instruments across epidemiologic studies makes cross-study comparisons and coherent causal determinations difficult, or whether these differences still produce interpretable findings for the field as a whole.^84^ We are well-positioned to further investigate this question of how different instruments pick up UFPs in future work. We observed, for example, a slightly non-linear relationship between the DiSCmini and all other PNC instruments when the predicted concentrations were high (SI Figure S21). A non-linear trend was also present when comparing the BC, NO_2_, PM_2.5_, and CO_2_ predictions to those from the DiSCmini, but less so when comparing these to the PNC predictions from other instruments. Furthermore, we will be able to use of size-resolved particle counts from the NanoScan (13 size bins, data not shown) or by looking at the differences between the unscreened and screened P-TRAKs, where the minimum sizes are 20 and 36 nm, respectively, in order to characterize size-specific exposure surfaces, sources, and health effects.

A feature of mobile monitoring campaigns is their reliance on repeated, short-term samples in order to achieve increased spatial coverage when compared to traditional long-term monitoring approaches. Since we collected about 29 two-minute samples per site (about an hour of data), we recognize that the resulting annual average site estimates are noisy. Still, with MSE-based R^2^ values of 0.77 for PNC and 0.60 for BC, our models performed better than many other short-term stationary and non-stationary monitoring campaigns (R^2^ of approximately 0.13-0.72 for PNC^21,42,43,49,50,53–56,58–63,68,85–91^ and 0.12-0.86 for BC.^21,53,58,63,68,71,72,75^ Figure 3 illustrates these results as well as those from other long-term stationary campaigns. There are several features of our study design that could have impacted our strong model performances. For PNC, Saha et al. (2019) reported that short-term stationary (collecting short-term samples while stopped, as opposed to while moving or traditional long-term stationary sampling) studies like ours have generally sampled between 60-644 sites, sampled each site between 15 minutes and 3 hours, and collected between 1-5 repeat samples per site. Similarly, BC studies like this one have generally sampled 26-161 sites, sampled each site about 30 minutes, and collected about 2-3 samples per site. Campaigns with more site counts have generally collected fewer repeat samples per site. Compared to earlier studies, we sampled more sites than most fixed and short-term stationary studies (309 sites). This dense monitoring network covered a larger geographic area and likely allowed us to capture hotspots that may have otherwise been missed by more sparse monitoring networks. Additionally, we visited each site for shorter periods of time (2 minutes), which allowed us to collect more repeat site visits (approximately 29) than what most studies have done. While our resulting total site sampling durations (∼58 minutes) were similar to other short-term stationary studies, we captured more temporal variability by sampling year-around during all days of the week and most times of the day, a limitation of most past campaigns. SI Figures S22-S23 summarize these as well as other short-term non-stationary mobile monitoring and long-term stationary designs for PNC^21,42,43,49,50,53–63,68,85–92^ and BC.^21,53,58,63–75^

**Figure 3.**
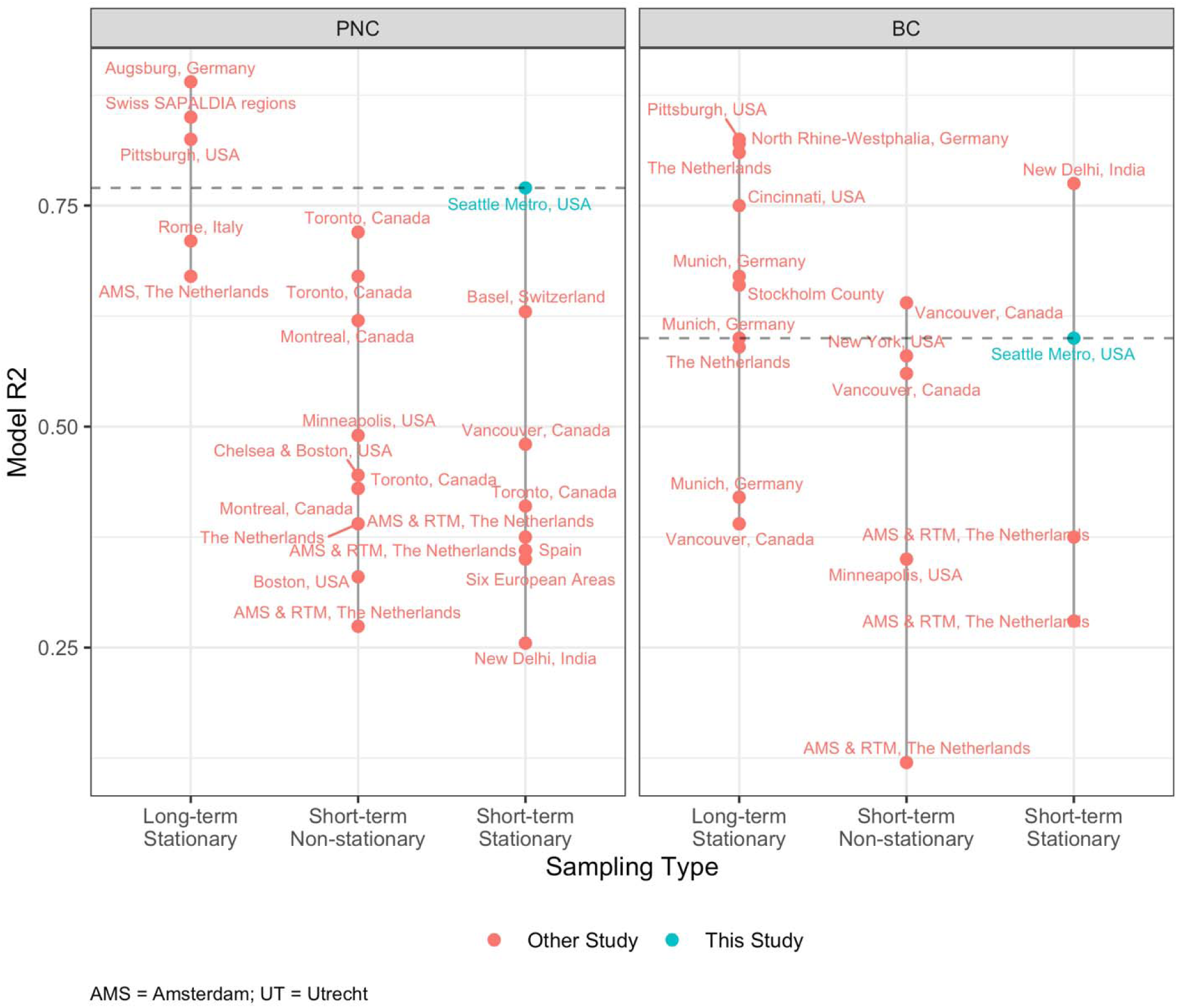
Cross-validated model R^2^ estimates from our and other PNC^21,42,43,49,50,53–63,68,85–90,92^ and BC^21,53,58,63–75^ studies. Studies are stratified by whether the sampling type was traditional, fixed site sampling (long-term stationary), short-term mobile monitoring campaigns that collected on-road data while in motion (short-term non-stationary), or short-term mobile monitoring campaigns that collected data while stopped (short-term stationary). Figure does not include Saha et al. (2021),^91^ who used a mixed sampling approach for PNC from multiple sources (R^2^ : 0.54-0.72). Horizontal dashed line is the R^2^ for this study. Plots show the average R^2^ from a study if multiple models were presented without a clear primary model.

In terms of our modeling approach, long-term averaging and winsorizing reduces the variability of the observations and focuses on the spatial contrasts of interest; this could have resulted in better performing models than had we modeled concentrations without aggregating them to a annual averages (e.g., stop medians). Sensitivity analyses using mean of (non-winsorized) medians, for example, generally resulted in slightly lower performing PNC and PM_2.5_ models due to the inclusion of more influential points in the models. Using a measure more robust to extreme observations, the median of medians, produced lower performing CO_2_ models due to the further reduction in variability. Still, we reported good out-of-sample MSE-based R^2^ estimates, which better characterize a model’s predictive performance at new locations and are generally lower than the in-sample regression-based R^2^ estimates that many studies report. We estimated these higher model performances despite the lower air pollution levels in our monitoring region, which can make it harder to get good prediction performance due to reduced variability (e.g., CO_2_).

Overall, these results demonstrates that the design of this campaign captured the spatial pollutant variations that can be explained by sensible land use features well, including those related to traffic. These data will thereby produce robust and representative long-term average TRAP exposures for the ACT cohort. Next steps include applying these prediction models to the cohort and conducting inferential analyses to determine the association of these pollutants with brain health. The rich dataset from this extensive campaign also provides an excellent foundation for investigating many important questions about how to best design mobile monitoring campaigns for application to subsequent epidemiologic studies.

## Data Availability

The data presented in this work may be available upon request.

## 5 Acknowledgements

We are grateful to our two drivers, Jim Sullivan and Dave Hardie, for all of their efforts collecting these data, and to Brian High for building and supporting the database.

This work was funded by the Adult Changes in Thought – Air Pollution (ACT-AP) Study (National Institute of Environmental Health Sciences [NIEHS], National Institute on Aging [NIA], R01ES026187), BEBTEH: Biostatistics, Epidemiologic & Bioinformatic Training in Environmental Health (NIEHS, T32ES015459), and the University of Washington Interdisciplinary Center for Exposure, Disease, Genomics & Environment (NIEHS, 2P30 ES007033-26). Research described in this article was conducted in part under contract to the Health Effects Institute (HEI), an organization jointly funded by the United States Environmental Protection Agency (EPA) (Assistance Award No. CR-83998101) and certain motor vehicle and engine manufacturers. The contents of this article do not necessarily reflect the views of HEI, or its sponsors, nor do they necessarily reflect the views and policies of the EPA or motor vehicle and engine manufacturers.

