## Supplementary Information for "Characterization of annual average traffic-related air pollution levels (particle number, black carbon, nitrogen dioxide, PM_2.5_, carbon dioxide) in the greater Seattle area from a year-long mobile monitoring campaign"

#### List of Figures

Figure S1. Covariate distributions of mobile monitoring stops (n=309) and ACT cohort locations (n=10,330). A1 and A2 roads are primary roads with and without limited access, respectively (e.g., interstate highways are A1, some US and state highways are A2). .......... 2
Figure S2. Mean percent error (estimated_conc – true_conc]/true_conc*100) in the estimated nitrogen monoxide (NO) and nitrogen dioxide ($NO_2$) annual average at the 10th & Weller (AQS10W; near-road site) and Beacon Hill (AQSBH; background site) regulatory sites from repeated short-term random samples (2- and 60-min), when compared to the "true" annual average estimated from all the available 2019 data. Estimates are for 10,000

Figure S12. Comparison of two-minute median concentrations from mobile monitoring and the Department of Ecology (DOE) readings at air quality system (AQS) collocation sites. MSE-based $R^2$: BC = 0.69, $NO_2$ = 0.71, $PM_{2.5}$ = 0.61. The dashed line is the 1-1 line; the blue line is the least squares linear regression fit. Mobile monitoring $PM_{2.5}$ concentrations are from calibrated nephelometer readings (see Methods). DOE $PM_{2.5}$ concentrations are from nephelometers when available (AQSD, AQSK, AQSTUK – readings are updated every minute), otherwise they are from gravimetric and beta attenuation (BAM) methods, which are updated less frequently (AQS10W – readings are based on rolling 1-hour estimates updated every 6 minutes, AQSBH – readings are updated hourly). ........ 29
Figure S22. Sampling approaches across our and other PNC studies.[46–69] Studies are stratified by whether the sampling type was traditional, fixed site sampling (long-term stationary), short-term mobile monitoring campaigns that collected on-road data while in motion (short-term non-stationary), or short-term mobile monitoring

campaigns that collected data while stopped (short-term stationary). Figure does not include Saha et al. (2021), who used a mixed sampling approach for PNC from multiple sources.[70] Note that little data were available for short-term non-stationary studies regarding visit duration, total site duration or visits per site. The single study under short-term non-stationary visit duration of ~ 8 min was conducted with pedestrians (Sabaliauskas et al. 2015). ..... 41
Figure S23 Sampling approaches across other BC studies.[46,51,56,61,62,71–81] Studies are stratified by whether the sampling type was traditional, fixed site sampling (long-term stationary), short-term mobile monitoring campaigns that collected on-road data while in motion (short-term non-stationary), or short-term mobile monitoring campaigns that collected data while stopped (short-term stationary). Note that little data were available for short-term non-stationary studies regarding visit duration, total site duration or visits per site. ..... 42

#### List of Equations

Equation S1. Nephelometer light scattering (bscat, m-1) calibration curve for $PM_{2.5}$. We fit this model using regulatory monitoring data between 1998-2017. Daily average measurements from nine non-industrial regulatory air monitoring sites in the region where both $PM_{2.5}$ (using federal reference methods) and nephelometer light scattering data were collected were used. We excluded the years 2008-2009 due to nephelometer instrumentation issues noted by the local regulatory agency. The model's leave-one-site-out cross-validated $R^2$ and root mean square error (RMSE) were 0.92 and 1.97 $\mu g/m^3$, respectively. ..... 8

#### List of Notes

### S1 Methods

#### S1.1 Study Design

*Note S1. Selecting the monitoring locations*

We used ArcMap to select 304 participant residences within our monitoring region that maximized spatial coverage. To do so, we first created a street network in ArcMap using Tiger/Line® shapefiles downloaded from the U.S. Census Bureau.[1,2] These include all roads within the monitoring area. We divided the monitoring area into nine regions and selected approximately 34 participant residences within each of these regions (~34 locations/region x 9 regions = 304 total locations). These locations were meant to maximize spatial coverage by minimizing the distance between each selected location and all the nearby participant locations. The initially selected locations were jittered (using the *jitter* function in R [v 3.5.1, using RStudio v 1.0.143]) to maintain participant confidentiality, and 304 new, nearby locations were identified as monitoring locations. The resulting locations were shifted anywhere from roughly a couple of houses to several blocks over. Locations were manually moved to the nearest home if the jittering caused locations to end up in a lake, park, etc. We used Google Maps Street View[3] to ensure that a vehicle could safely park at each location, otherwise the stop was moved to the nearest location where it was safe to do so. We included 5 regulatory monitoring locations to obtain the final 309 monitoring locations.

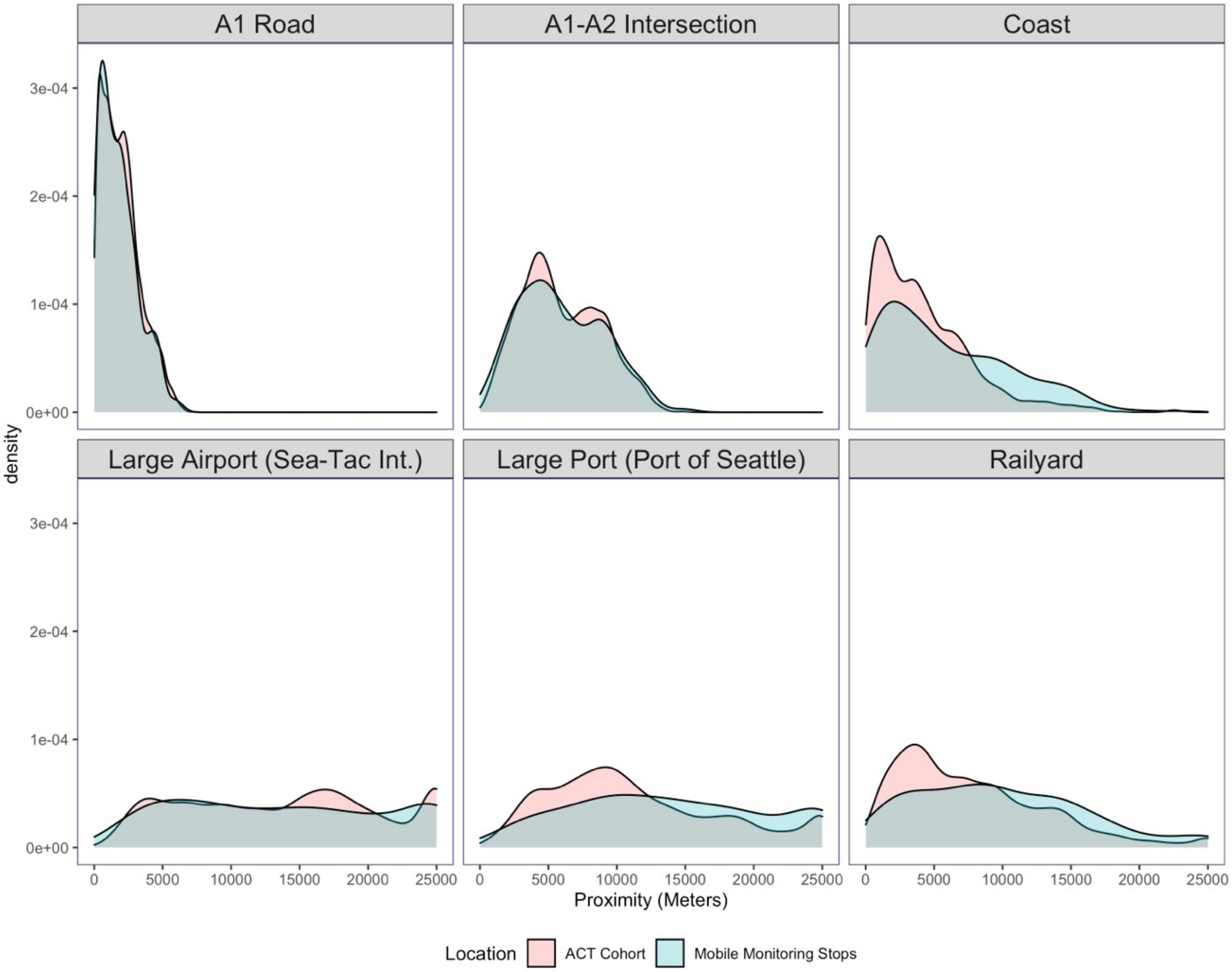

*Figure S1. Covariate distributions of mobile monitoring stops (n=309) and ACT cohort locations (n=10,330). A1 and A2 roads are primary roads with and without limited access, respectively (e.g., interstate highways are A1, some US and state highways are A2).*

*Table S1. Route Statistics.*

| Route[a] | No. Stops | No. Sampling Dates | Distance (mi) | Distance (km) | Total Distance (mi)[b] | Total Distance (km) | Median (IQR) Drive Time (hr) | Total Drive Time (hr) |
|---|---|---|---|---|---|---|---|---|
| 1 | 40 | 30 | 48 | 78 | 1,446 | 2,328 | 5.2 (4.8, 5.7) | 153 |
| 2 | 35 | 30 | 47 | 75 | 1,397 | 2,247 | 4.8 (4.4, 5.1) | 140 |
| 3 | 45 | 34 | 59 | 95 | 2,008 | 3,232 | 5.6 (4.9, 6.1) | 170 |
| 4 | 33 | 33 | 66 | 107 | 2,190 | 3,525 | 5.0 (4.7, 5.3) | 143 |
| 5 | 32 | 32 | 69 | 112 | 2,221 | 3,574 | 5.1 (4.7, 5.5) | 143 |
| 6 | 35 | 32 | 88 | 142 | 2,829 | 4,554 | 5.4 (4.9, 5.8) | 141 |
| 7 | 32 | 30 | 90 | 145 | 2,697 | 4,341 | 5.6 (5.2, 6.1) | 163 |
| 8 | 28 | 32 | 104 | 168 | 3,332 | 5,362 | 5.7 (5.2, 6.1) | 156 |
| 9 | 29 | 35 | 92 | 148 | 3,227 | 5,193 | 5.5 (5.0, 5.8) | 158 |
| Total | 309 | 35 | 664 | 1,069 | 21,347 | 34,355 | 5.2 (4.8, 5.8) | 1,367 |

[a] There were about 18 additional make-up drives (about 4-5 per quarter), each with stops from multiple routes.

[b] Total route driving distance is estimated from the route distance and the number of sampling dates. The exact distance varied based on make-up drive, route deviations, etc.

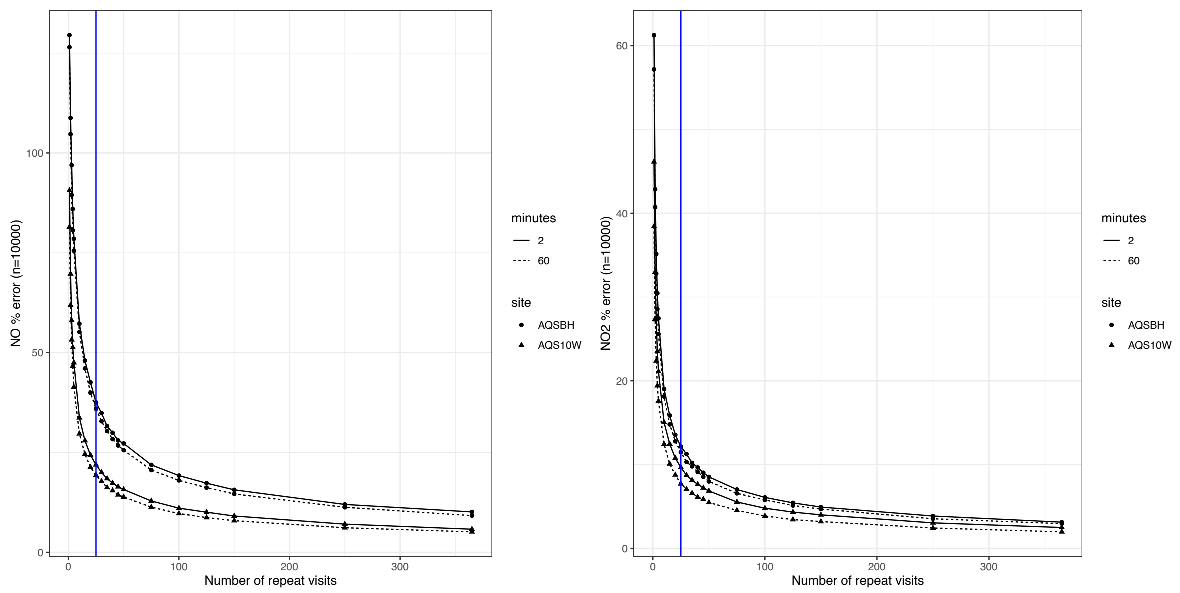

*Figure S2. Mean percent error (estimated_conc – true_conc]/true_conc*100) in the estimated nitrogen monoxide (NO) and nitrogen dioxide ($NO_2$) annual average at the 10th & Weller (AQS10W; near-road site) and Beacon Hill (AQSBH; background site) regulatory sites from repeated short-term random samples (2- and 60-min), when compared to the "true" annual average estimated from all the available 2019 data. Estimates are for 10,000 simulations of random sampling without replacement. The blue vertical line is for 25 repeat visits. NO was used in this simulation because it is a quick decaying pollutant representative of various other TRAPs (e.g., PNC, BC). $NO_2$ is what we measured in or campaign and a slower decaying pollutant.*

*Table S2. Air pollutants and other parameters measured with mobile monitoring.*

| Parameter | Instrument | Manufacturer | Measurement Range | Limit of Quantification | Time Resolution |
|---|---|---|---|---|---|
| **Particles (pt)** | | | | | |
| PNC[a] | | | | | |
| 10-420 nm (13-bin PSD[a]) | NanoScan 3910 | TSI | $10^2$-$10^6$ pt/cm$^3$ | 10 pt/ cm$^3$ | 60 sec |
| 10-700 nm | DiSCmini | Testo | $10^3$-$0^6$ pt/cm$^3$ | 500-2,000 pt/cm$^3$ [b] | 1 sec |
| 20-1,000 nm | PTRAK 8525 | TSI | 0-5x$10^5$ pt/cm$^3$ | 1 pt/cm$^3$ | 1 sec |
| 36-1,000 nm | PTRAK 8525, with diffusion screen | TSI | 0-5x$10^5$ pt/cm$^3$ | 1 pt/cm$^3$ | 1 sec |
| BC | microAeth MA200 | AethLabs | 0-$10^6$ ng/m$^3$ | 30 ng BC/m$^3$ [c] | 10 sec |
| Light scattering nephelometer ($PM_{2.5}$) | M903 | Radiance Research | 0 - >1 km$^{-1}$ | $10^{-6}$ m$^{-1}$ | 10 sec |
| **Gases** | | | | | |
| $NO_2$ | CAPS $NO_2$ | Aerodyne Research, Inc. | 0-2x$10^3$ ppb | 2 ppbv | 1 sec |
| $CO_2$ | LI-850 | Li-Cor | 0-5x$10^3$ ppm (vol) | 100 ppmv | 1 sec |
| CO[d] | CO Monitor T15N | Langan, Inc. | 0-200 ppm | 0.1 ppm | 1 sec |
| **Other** | | | | | |

| | | | | | |
|---|---|---|---|---|---|
| Temperature | Onset UX100-011 | HOBO | -4-158°F | | 1 sec |
| Relative humidity | Onset UX100-011 | HOBO | 0-95% | | 1 sec |
| Positioning & real-time tracking | DG-500 | US GlobalSat | 0-515 m/sec speed | 2.5 m | 1 sec |

[a] PNC: particle number concentration; PSD: particle size distribution

[b] estimate; detection limit is dependent on particle size

[c] for a 5 min time base, 150 ml/min flow rate

[d] CO measurements were collected but not utilized because they did not meet our quality assurance standards

Instruments were in the back of the vehicle where they were powered by two rechargeable batteries and connected to gas- or particle- specific manifolds (SI Figure S3-S4). These were connected to a rooftop inlet facing the front of the car to reduce the possibility of self-contamination while in motion. Instrument clocks were all synchronized at the beginning of each drive within ~2 seconds. Instruments were started within an hour before the start of each drive and continuously run until the end of the route.

Drivers were instructed to follow the specific Google Map route directions and to take notes of any field anomalies (e.g., sampling behind a school bus or next to a construction site).

Instrument data files were uploaded to a secure data management system (MySQL) after each drive using standardized naming conventions to automate data uploads and the generation of daily data reports (see below for further details).

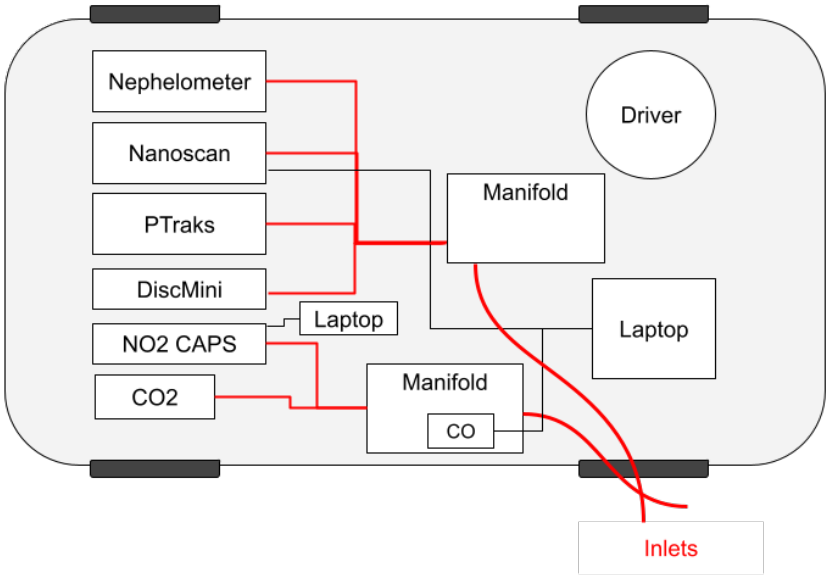

*Figure S3. In-vehicle configuration*

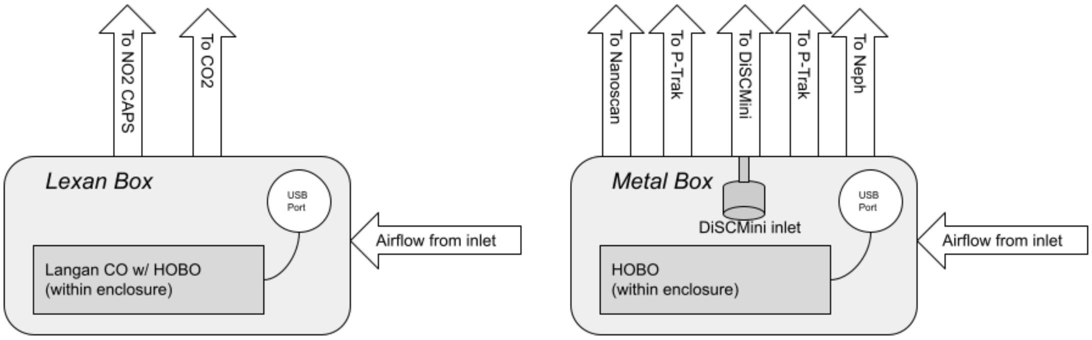

*Figure S4. Manifold schematics*

#### S1.2 Computation

*Note S3. Software used in analyses.*

We conducted all analyses using MySQL[4] and R (v 3.6.2, using RStudio v 1.2.5033).[5] We used the R packages: Broom (v. 0.5.5),[6] colorspace (v. 1.4-1),[7] cowplot (v. 1.0.0),[8] dplyr (v. 1.0.6),[9] fmsb (v. 0.7.1),[10] forcats (v. 0.5.0),[11] GGally (v. 2.1.1),[12] ggmap (v. 3.0.0),[13] ggplot2 (v. 3.3.3),[14(p2)] ggpmisc (v. 0.4.0),[15] ggpp (v. 0.4.0),[16] ggpubr (v. 0.2.5),[17] ggrepel (v. 0.8.1),[18] ggspatial (v. 1.1.4),[19] gstat (v. 2.0-7),[20] kableExtra (v. 1.1.0),[21] knitr (v. 1.28),[22] lubridate (v. 1.7.10),[23] magrittr (v. 1.5),[24] pls (v. 0.0.1),[25] purrr (v. 0.3.3),[26] readr (v. 1.3.1),[27] sf (v. 0.9-5),[28] spData (v. 0.3.10),[29] stringr (v. 1.4.0),[30] tibble (v. 3.1.2),[31] tidyr (v. 1.0.2),[32] tidyverse (v. 1.3.0),[33] units (v. 0.6-7)[34] and VCA (v. 1.4.2).[35] We created all maps with map tiles by Stamen Design[36] under CC BY 3.0,[37] using data by OpenStreetMap under ODbL.[38]

*Equation S1. Nephelometer light scattering (bscat, m-1) calibration curve for $PM_{2.5}$. We fit this model using regulatory monitoring data between 1998-2017. Daily average measurements from nine non-industrial regulatory air monitoring sites in the region where both $PM_{2.5}$ (using federal reference methods) and nephelometer light scattering data were collected were used. We excluded the years 2008-2009 due to nephelometer instrumentation issues noted by the local regulatory agency. The model's leave-one-site-out cross-validated $R^2$ and root mean square error (RMSE) were 0.92 and 1.97 µg/m³, respectively.*

$$PM_2\left(\frac{ug}{m^3}\right) = 25.10x10^4(bscat) + 1.06$$

#### S1.3 Quality Assurance and Quality Control

*Note S4. Quality assurance and quality control procedures*

##### *Data Management System*

After each drive, the field technician uploaded each instrument's file to the server using a standardized file naming convention. These files were automatically loaded into a MySQL database every morning at 4 a.m. A report was produced automatically that showed the time series plot of each instrument, counts of the times of day that each stop on the route had been visited to date, and a map that highlighted any missed stops. The driver reviewed these data before starting the next day's drive. Single missed stops could be visited on the way to another day's route or on a day dedicated to make-up stops. In addition, a project manager, information technology (IT) specialist, and several data analysts routinely reviewed and worked with the data, thus allowing for additional feedback.

We carried out an extensive independent code review of the database and made further improvements to the system before completely reloading all raw data files into the database and locking the final version.

##### *Instrumentation*

To ensure instrument accuracy, all gas instruments were calibrated in our laboratory before the campaign and every few weeks thereafter. Particle instruments were purchased new and arrived with calibration certifications, or they were compared to like instruments that had been serviced prior to the study. Primary and backup instruments were collocated every few weeks on route to assess the precision (repeatability) of our measurements in different environments and over time.

##### *Data Cleaning*

We conducted various quality control procedures prior to conducting data analyses. We added a ten-second lag to all the instrument readings to account for the time required for a volume of air to travel from the sampling inlet to each instrument. This was based on the manifold volumes and instrument flow rates.

Readings with instrument error codes were dropped. This included, for example, aethalometer (BC) pump flow errors and readings of $NO_2$ field baseline samples.

Aethalometers were checked to ensure that the filter attenuation was below 50% thus ensuring optimal instrument sensitivity at all times.[39,40]

Gas instruments and nephelometers (which were checked for a response against $CO_2$ gas[41]) were calibrated various times during the study period. CO and nephelometer instruments were automatically reset during calibrations. $CO_2$ and $NO_2$ instruments were manually calibrated using least squares linear regression models with reference concentrations as the independent variable and instrument readings as the response variable.[42,43] Particle instruments were checked for zero concentration responses by placing a high efficiency particle air (HEPA) filter on the instrument or manifold inlet.

We calculated stop visit medians (from about 2 minutes worth of data). Readings outside the instrument ranges, screened P-TRAK readings below 100 pt/cm$^3$, and other PNC instrument readings below 300 pt/cm$^3$ (NanoScans, unscreened P-TRAKS) were dropped.

We investigated collocated instrument readings to assess repeatability. Comparing instruments to one another is particularly common with particle instruments since there is no standard for the field calibration of these instruments. Backup $NO_2$ ("NO2_1") and NanoScan ("PMSCAN_3") instruments were adjusted based on readings from their respective primary instruments during the beginning of the study since these were used exclusively at the beginning of the study. We calibrated PNC readings from the two DiSCmini instruments used in this study to the mean of their responses.[44] This was done to ensure consistency across instruments since these were equally used throughout the study period. In this approach, a calibration curve is established by fitting separate linear regression models to each instrument, with that instrument's readings as the independent variable and the mean reading of duplicate instruments as the response variable.

The backup $CO_2$ instrument (CO2_19) was dropped since it produced unstable responses over time, it did not always correlate well with the primary instrument (CO2_14), and it was solely used as a collocation instrument (i.e., never on its own). All CO readings were dropped since instruments produced unstable readings, and collocated instruments were poorly correlated with one another or observations from collocations at regulatory monitoring sites.

*Quality Control Results Summary*

SI Table S3 shows the calibration curve coefficient estimates used to manually adjust CO and $NO_2$.

In response to clean, filtered air, particle instruments generally reported near zero concentrations that were also lower than a “low” ambient concentration, as determined from the data (SI Figure S5). Some exceptions included the backup aethalometer (BC_0066), which reported negative readings, though this was based on very little data (two 2-min medians). The primary aethalometer (BC_0063) and nephelometer instruments (PM25_176), as well as the backup DiSCmini instrument (PMDISC_8) additionally reported low ambient concentrations that were similar to some of their filtered air responses, suggesting that these instruments may be less sensitive to very low ambient concentrations.

Collocated instruments generally produced similar responses (SI Figure S6). As noted above, the backup $CO_2$ instrument (CO2_19) and all CO instruments were dropped because they did not meet quality assurance standards. Backup $NO_2$ and NanoScan instruments (NO2_1, PMSCAN_3) were adjusted to better align with primary instrument readings.

Temperature and relative humidity conditions inside the manifold during site visits are presented in SI Table S4.

*Table S3. Distribution of calibration curve coefficient estimates.*

| Pollutant[a] | Instrument ID | Term | N[b] | Min | Median | Max |
|---|---|---|---|---|---|---|
| $NO_2$ (ppb) | NO2_1 | slope | 6 | 1.09 | 1.14 | 1.20 |
| $NO_2$ (ppb) | NO2_1 | intercept | 6 | -29.02 | -3.15 | -0.55 |
| $NO_2$ (ppb) | NO2_2 | slope | 18 | 0.55 | 1.05 | 1.23 |
| $NO_2$ (ppb) | NO2_2[c] | intercept | 18 | 0 | 0 | 0 |
| CO (ppm) | CO_1 | slope | 16 | 1.05 | 1.10 | 1.16 |
| CO (ppm) | CO_1 | intercept | 16 | 1.60 | 2.25 | 2.77 |
| CO (ppm) | CO_190134 | slope | 5 | 0.74 | 0.84 | 0.94 |
| CO (ppm) | CO_190134 | intercept | 5 | 1.46 | 2.19 | 2.27 |
| CO (ppm) | CO_3 | slope | 13 | 0.20 | 0.41 | 0.82 |
| CO (ppm) | CO_3 | intercept | 13 | 0.81 | 1.33 | 2.29 |

[a] $CO_2$ was automatically reset after each calibration, and no additional adjustments were necessary.

[b] N = number of calibration days.

[c] A no intercept model was fit to instrument NO2_2, which reset after each baseline zero reading.

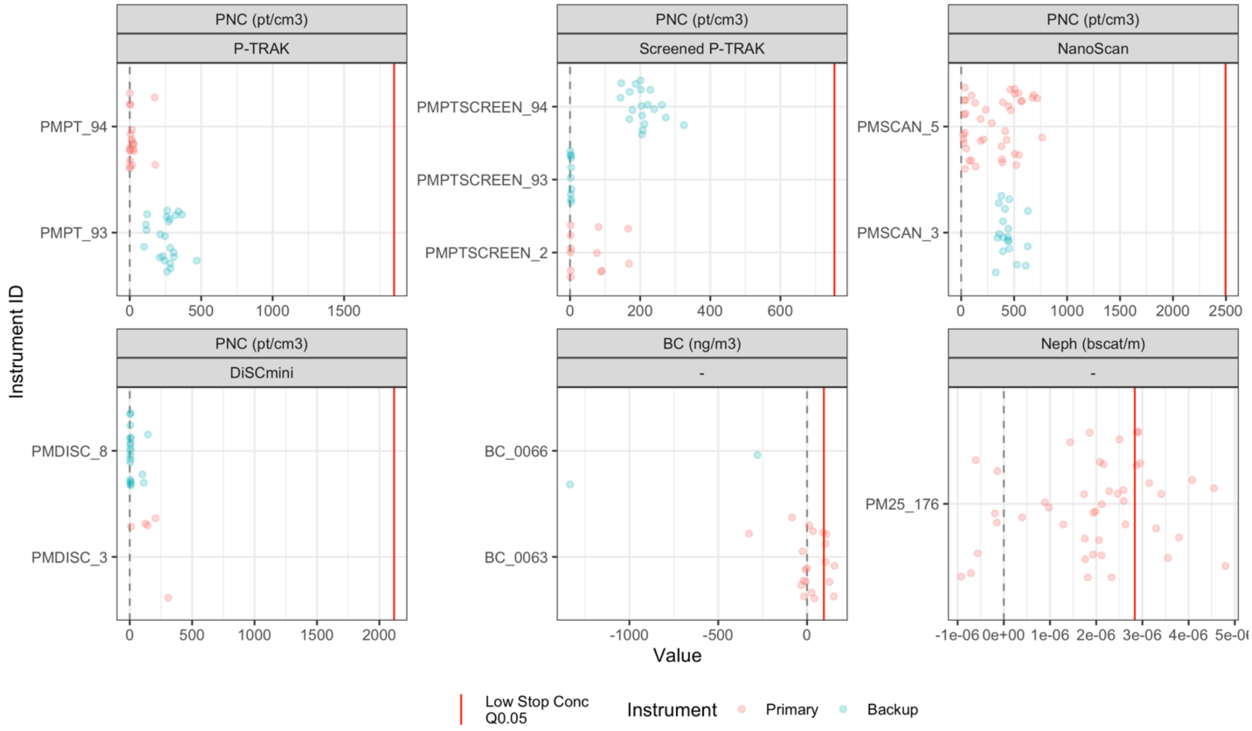

*Figure S5. Particle instrument (y-axis) responses to filtered air concentrations (x-axis; near 0 pt/cm³). Dots show median, two-minute instrument readings. Red lines are "low" ambient concentration references, based on the 5th quantile of stop concentrations for each pollutant.*

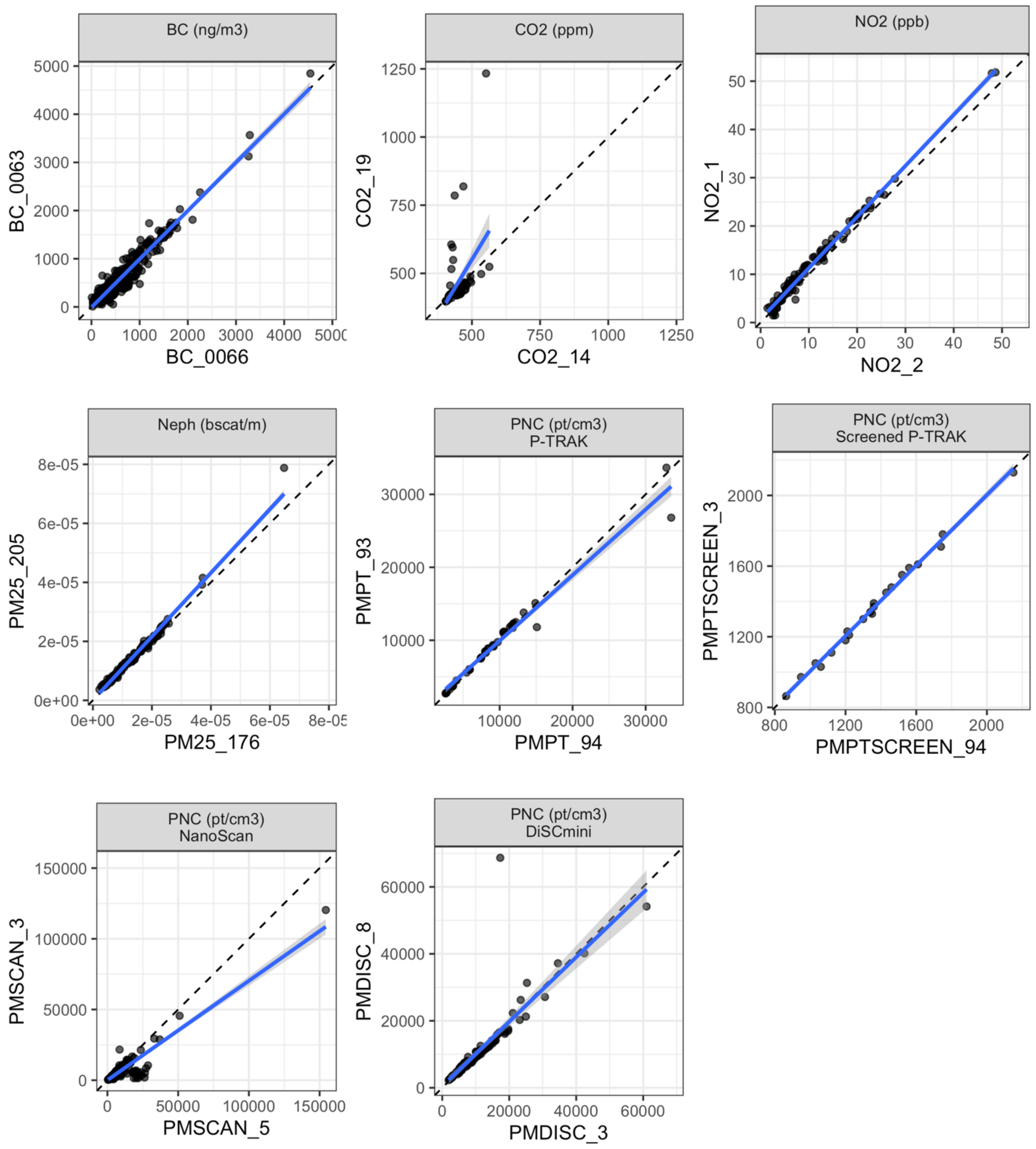

*Figure S6. Comparison of two-minute median stop concentrations from instrument collocations. Gas values are post calibration.*

*Table S4. Distribution of temperature and relative humidity conditions inside the manifold during site visits (N=9,047 total). a*

| Variable | Min | Q05 | Q25 | Median | Q75 | Q95 | Max |
|---|---|---|---|---|---|---|---|
| Relative Humidity (%) | 12 | 26 | 37 | 44 | 51 | 61 | 78 |
| Temperature (F) | 50 | 60 | 64 | 67 | 72 | 80 | 96 |

[a] Measurements are for 2-min medians from within the vehicle (manifold) for 9,047 site visits

*Table S5. Collocation regulatory sites and similar parameters measured.a*

| Station (ID) | Location | $PM_{2.5}$ FRM | $PM_{2.5}$ FEM | bscat & $PM_{2.5}$ bscat | BC | $NO_2$[b] |
|---|---|---|---|---|---|---|
| $10^{th}$ & Weller, Seattle (BK) | Urban Center; near-road | No | Yes | No | Yes | Yes |
| Tukwila Allentown (BL) | Suburban, industrial, residential | No | Yes | Yes | Yes | No |
| Beacon Hill (BW) | Suburban, commercial, residential | Yes | Yes | No | No | Yes |
| Duwamish (CE) | Urban center, industrial | No | Yes | Yes | Yes | No |
| James St & Central Ave, Kent (CW) | Suburban, commercial | No | Yes | Yes | Yes | No |

[a] FRM: federal reference method; FEM: federal equivalent method; bscat: beta light scattering; Temp: temperature (˚F); RH: relative humidity

[b] or $NO_y$-NO

#### S1.4 Prediction Models

*Note S5. Geographic covariates*

There were 350 initial geographic covariates (geocovariates) that were reduced to 191 taking a similar approach as past work (SI Table S6).[19,20,45] Using the training-validation data set (90%, n=278 sites), first, we excluded variables if they lacked variability (less than 40% of the data were different from the most common value) since these were not likely to improve, and could even worsen, the model fit. Next, we excluded all land use proportion variables where the maximum proportion observed in the data was less than 20%. Low values for these variables indicated that these land use types made up a small fraction of the land relative to other land use variables and would not likely have a meaningful impact on observed pollutant concentrations. We dropped variables if too many outliers were observed in the data (>2% of the total data). Finally, we log-transformed proximity variables to better model pollutant exponential decay with increasing source distance.

*Table S6. Available geographic covariates (geocovariates) used in UK-PLS models.*

| Kind | Covariate | Buffers | Description |
|---|---|---|---|
| airports | log_m_to_airp | 0 | log meters to closest airport |
| airports | log_m_to_l_airp | 0 | log meters to closest large airport |
| bus | bus_s | 100, 150, 300, 400, 500, 750, 1000, 1500, 3000, 5000 | sum of bus routes |
| bus | log_m_to_bus | 0 | log meters to bus route |
| coast | log_m_to_coast | 0 | log meters to closest coastline |
| columnar NO2 | no2_behr | 0 | columnar NO2, mean from 2005-2007 |
| commercial and services | log_m_to_comm | 0 | log meters to closest commercial and services area |
| elevation | elev_above | 1000, 5000 | number of points (out of 24) more than 20 m and 50 m uphill of a location for a 1000 m and 5000 m buffer, respectively |
| elevation | elev_at_elev | 1000, 5000 | number of points (out of 24) within 20 m and 50 m of the location' elevation for a 1000 m and 5000 m buffer, respectively |
| elevation | elev_below | 1000, 5000 | number of points (out of 24) more than 20 m and 50 m downhill of a location for a 1000 m and 5000 m buffer, respectively |
| elevation | elev_elevation | 0 | elevation above sea level in meters |

| | | | |
|---|---|---|---|
| elevation | elev_stdev | 1000, 5000 | standard deviation of elevation of 20 points surrounding the location |
| imperviousness | imp_a | 50, 100, 150, 300, 400, 500, 750, 1000, 3000, 5000 | average imperviousness |
| land use | rlu_decid_forest_p | 500, 750, 1000 | proportion of deciduous forest |
| land use | rlu_dev_hi_p | 300, 400, 500, 750, 1000, 3000, 5000 | proportion of highly developed land (e.g., commercial and services; industrial; transportation, communication and utilities) |
| land use | rlu_dev_lo_p | 50, 100, 150, 300, 400, 500, 750, 1000, 3000, 5000 | proportion of low developed land (e.g., residential) |
| land use | rlu_dev_med_p | 50, 100, 150, 300, 400, 500, 750, 1000, 3000, 5000 | proportion of medium developed land (e.g., residential) |
| land use | rlu_dev_open_p | 150, 300, 400, 500, 750, 1000, 3000, 5000 | proportion of developed open land |
| land use | rlu_evergreen_p | 400, 500, 750, 1000 | proportion of evergreen forest |
| land use | rlu_mix_forest_p | 500, 750, 1000, 5000 | proportion of mixed forest |
| NDVI | ndvi_q25_a | 250, 500, 1000, 2500, 5000, 7500, 10000 | NDVI (25th quantile) |
| NDVI | ndvi_q50_a | 250, 500, 1000, 2500, 5000, 7500, 10000 | NDVI (50th quantile) |
| NDVI | ndvi_q75_a | 250, 500, 1000, 2500, 5000, 7500, 10000 | NDVI (75th quantile) |

| | | | |
|---|---|---|---|
| NDVI | ndvi_summer_a | 250, 500, 1000, 2500, 5000, 7500, 10000 | average summer time NDVI |
| NDVI | ndvi_winter_a | 250, 500, 1000, 2500, 5000, 7500, 10000 | average winter time NDVI |
| population | pop10_s | 500, 1000, 1500, 2000, 2500, 3000, 5000, 10000, 15000 | 2010 population density |
| port | log_m_to_l_port | 0 | log meters to closest large port |
| railroads, rail yards | log_m_to_rr | 0 | log meters to closest railroad |
| railroads, rail yards | log_m_to_ry | 0 | log meters to closest rail yard |
| roads | intersect_a1_a3_s | 3000 | number of A1-A3 road intersections |
| roads | intersect_a3_a3_s | 500, 1000, 3000 | number of A3-A3 road intersections |
| roads | ll_a1_s | 1500, 3000, 5000 | length of A1 roads |
| roads | ll_a2_s | 5000 | length of A2 roads |
| roads | ll_a23_s | 100, 150, 300, 400, 500, 750, 1000, 1500, 3000, 5000 | length of A2 and A3 roads |
| roads | ll_a3_s | 100, 150, 300, 400, 500, 750, 1000, 1500, 3000, 5000 | length of A3 roads |
| roads | log_m_to_a1 | 0 | log meters to closest A1 road |
| roads | log_m_to_a1_a1_intersect | 0 | log meters to closest A1-A1 road intersection |

| | | | |
|---|---|---|---|
| roads | log_m_to_a1_a2_intersect | 0 | log meters to closest A1-A2 road intersection |
| roads | log_m_to_a1_a3_intersect | 0 | log meters to closest A1-A3 road intersection |
| roads | log_m_to_a123 | 0 | log meters to closest A1, A2 or A3 road |
| roads | log_m_to_a2 | 0 | log meters to closest A2 road |
| roads | log_m_to_a2_a2_intersect | 0 | log meters to closest A2-A2 road intersection |
| roads | log_m_to_a2_a3_intersect | 0 | log meters to closest A2-A3 road intersection |
| roads | log_m_to_a23 | 0 | log meters to closest A2 or A3 road |
| roads | log_m_to_a3 | 0 | log meters to closest A3 road |
| roads | log_m_to_a3_a3_intersect | 0 | log meters to closest A3-A3 road intersection |
| stack emissions | em_CO_s | 3000, 15000, 30000 | sum of CO stack emissions |
| stack emissions | em_NOx_s | 15000, 30000 | sum of NOx stack emissions |
| stack emissions | em_PM10_s | 15000, 30000 | sum of PM10 stack emissions |
| stack emissions | em_PM25_s | 15000, 30000 | sum of PM2.5 stack emissions |
| stack emissions | em_SO2_s | 15000 | sum of SO2 stack emissions |
| truck routes | log_m_to_truck | 0 | log meters to closest truck route |
| truck routes | tl_s | 750, 1000, 1500, 3000, 5000, 10000, 15000 | length of truck routes |

| | | | |
|---|---|---|---|
| water | log_m_to_waterway | 0 | log meters to closest waterway |
| water | rlu_water_p | 1000, 3000, 5000 | proportion of water |

### S2 Results

#### S2.1 Site Visits

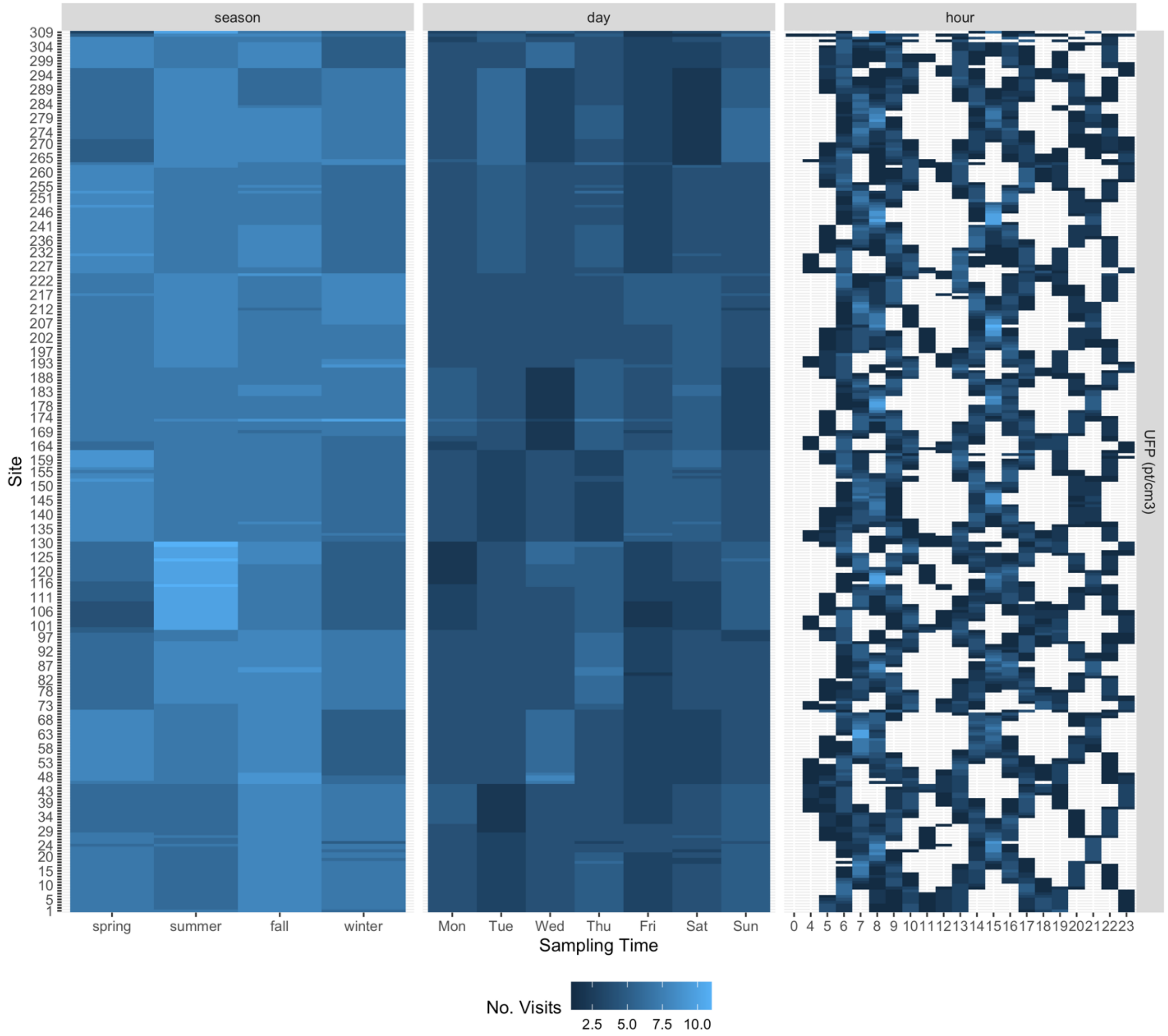

*Figure S7. Number of site visits per time period. Showing PNC data, though all instruments were similar.*

*Table S7. Original and final mobile monitoring stop measurements (~2 min each).*

| Pollutant[a] | Original Stop Measurements[b] | | Dropped Stop Measurements[c] | | Final Stop Measurements[b] | |
|---|---|---|---|---|---|---|
| | N | % | N | % | N | % |
| $CO_2$ (ppm) | 8, 982 | 99.28% | 32 | 0.36% | 8, 950 | 98.93% |
| BC ($ng/m^3$) | 9, 005 | 99.54% | 144 | 1.6% | 8, 861 | 97.94% |
| Neph (bscat/m) | 8, 802 | 97.29% | 16 | 0.18% | 8, 786 | 97.12% |
| $NO_2$ (ppb) | 8, 913 | 98.52% | 147 | 1.65% | 8, 766 | 96.89% |
| PNC ($pt/cm^3$), P-TRAK | 8, 731 | 96.51% | 2 | 0.02% | 8, 729 | 96.49% |
| PNC ($pt/cm^3$), screened P-TRAK | 8, 908 | 98.46% | 0 | 0% | 8, 908 | 98.46% |
| PNC ($pt/cm^3$), NanoScan | 9, 000 | 99.48% | 1 | 0.01% | 8, 999 | 99.47% |
| PNC ($pt/cm^3$), DiSCmini | 8, 790 | 97.16% | 93 | 1.06% | 8, 697 | 96.13% |
| TOTAL | 71, 131 | 98.28% | 435 | 0.61% | 70, 696 | 97.68% |

[a] PNC units are particles (pt) per $cm^3$.

[b] Original and final stop measurement percents are based on the total number of stops that collected at least one 2-minute measurement in the campaign (9,047; Total = 72,376 = 9,047 stops x 8 instruments).

[c] Measurements were dropped for various reasons: readings outside of each instrument's reporting range; NanoScan and non-screened P-TRAK readings < 300 $pt/cm^3$; backup $CO_2$

instrument (CO2_19) and all CO (both instruments) readings because these did not meet QC protocols (see Note S4). Dropped stops percents are based on the original stop measurements (the measurements actually collected).

*Table S8. Distribution of winsorized median site visit concentrations (N = 309 sites x ~ 29 visits/site).*

| **Pollutant** | | **N** | **Q05** | **Q25** | **Median** | **Mean** | **Q75** | **Q95** |
|---|---|---|---|---|---|---|---|---|
| PNC (pt/cm$^3$) | P-TRAK | 8,728 | 1,850 | 3,640 | 5,850 | 7,454 | 9,131 | 18,032 |
| PNC (pt/cm$^3$) | Screened P-TRAK | 8,908 | 754 | 1,580 | 2,520 | 3,285 | 4,050 | 8,136 |
| PNC (pt/cm$^3$) | NanoScan | 8,999 | 2,496 | 5,060 | 8,150 | 10,762 | 13,165 | 27,235 |
| PNC (pt/cm$^3$) | DiSCmini | 8,697 | 2,118 | 4,336 | 7,028 | 9,889 | 11,413 | 24,575 |
| BC (ng/m$^3$) | | 8,860 | 94 | 242 | 402 | 584 | 694 | 1,736 |
| $NO_2$ (ppb) | | 8,747 | 1.7 | 4.1 | 7.4 | 9.4 | 13 | 24 |
| $PM_{2.5}$ (µg/m$^3$) | | 8,786 | 1.8 | 2.7 | 3.9 | 4.8 | 5.8 | 11 |
| $CO_2$ (ppm) | | 8,950 | 405 | 415 | 425 | 431 | 441 | 478 |

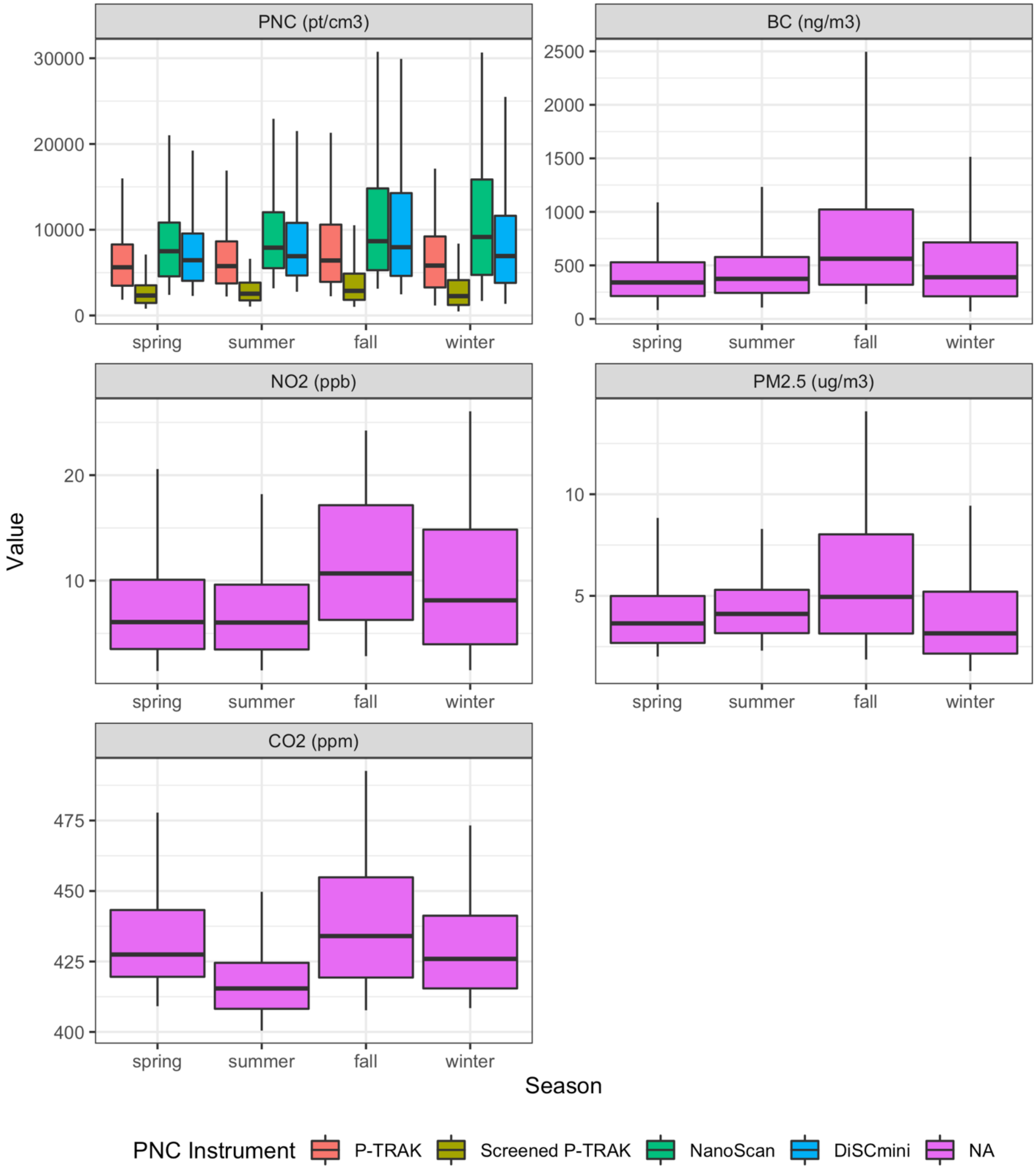

*Figure S8. Distribution of winsorized median site visit concentrations by season. Boxes show the 25th, 50th and 75th quantiles; whiskers show the 5th and 95th quantiles. The "NA" PNC legend value refers to pollutants other than PNC.*

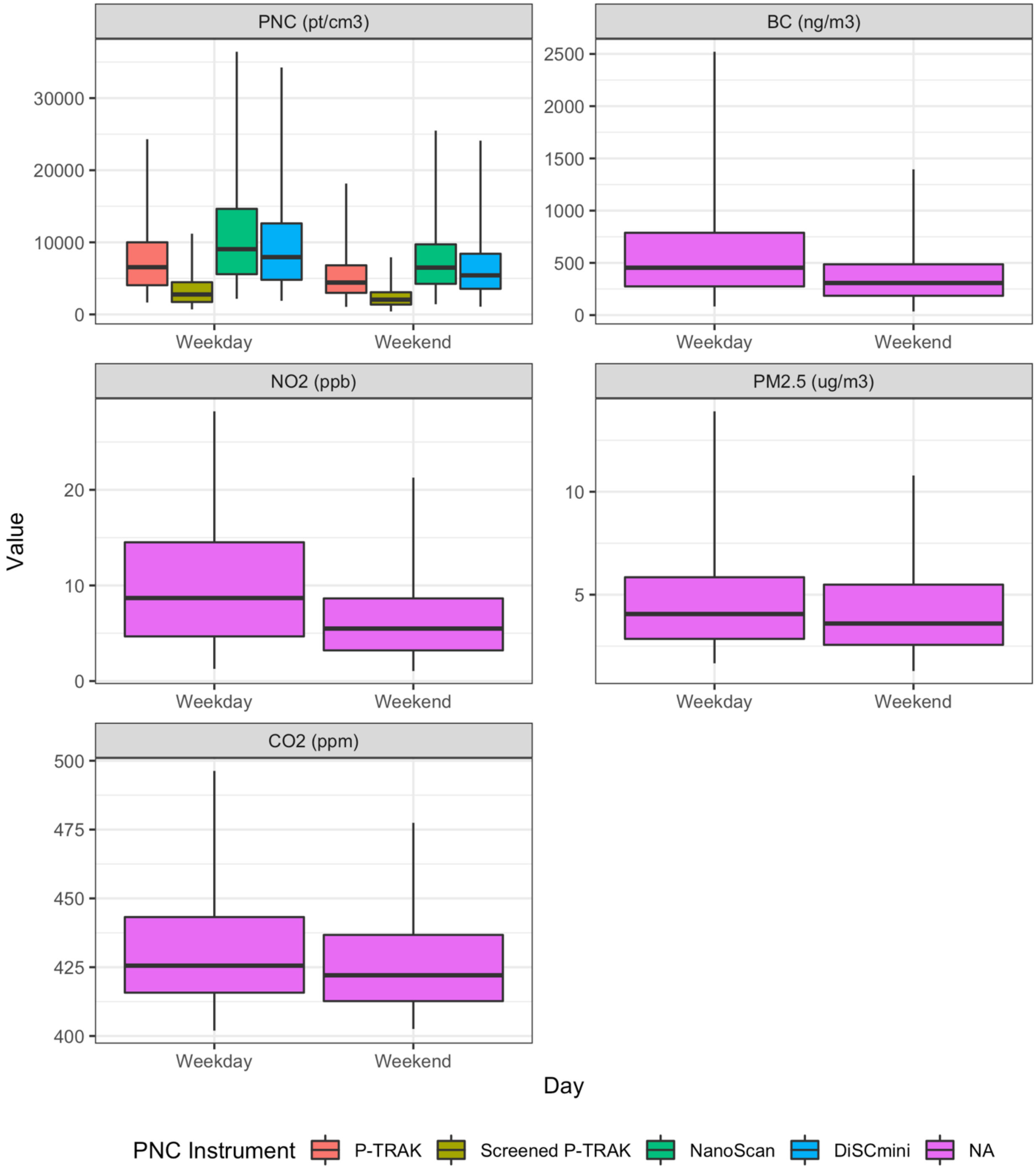

*Figure S9. Distribution of winsorized median site visit concentrations by day of the week. Boxes show the 25th, 50th and 75th quantiles; whiskers show the 5th and 95th quantiles. The "NA" PNC legend value refers to pollutants other than PNC.*

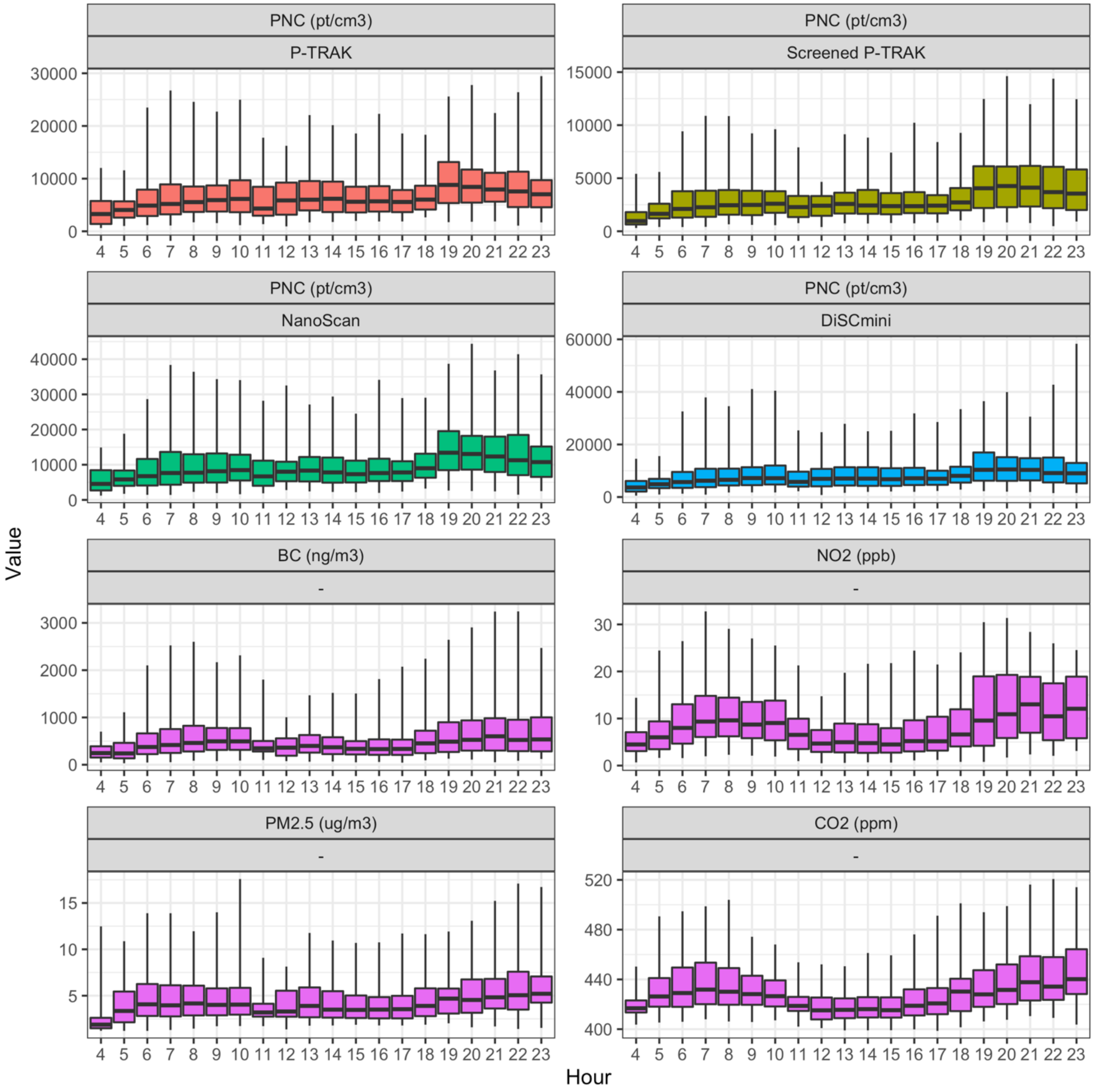

*Figure S10. Distribution of winsorized median site visit concentrations by hour of the day. Boxes show the 25th, 50th and 75th quantiles; whiskers show the 5th and 95th quantiles.*

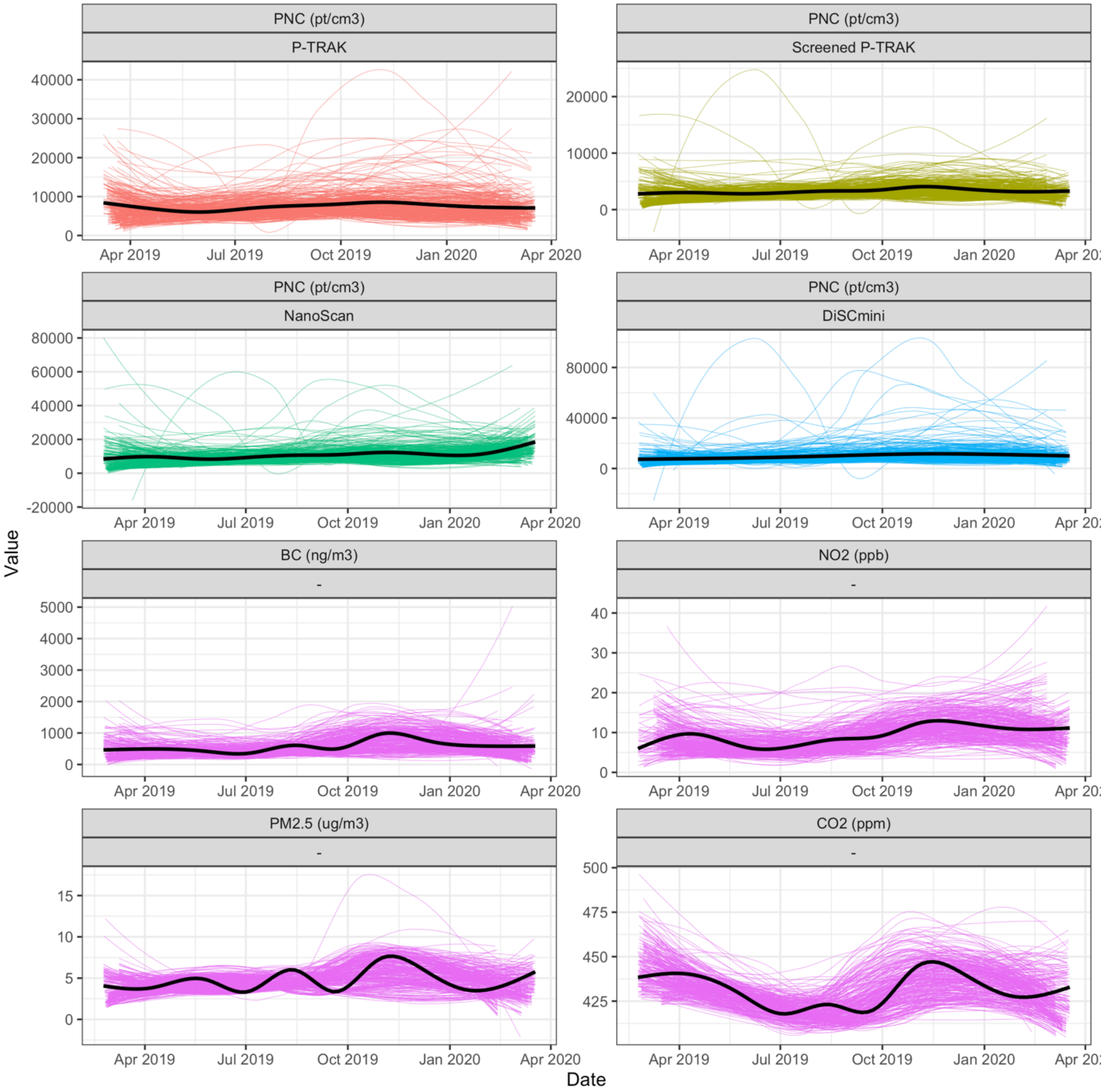

*Figure S11. Site-specific concentrations over the course of the study. Thin lines show site-specific smooth (loess) fits for winsorized median visit concentrations (N~29 visits/site). Black lines show the overall smooth trends for all the sites.*

#### S2.2 Collocations at Regulatory Monitoring Sites

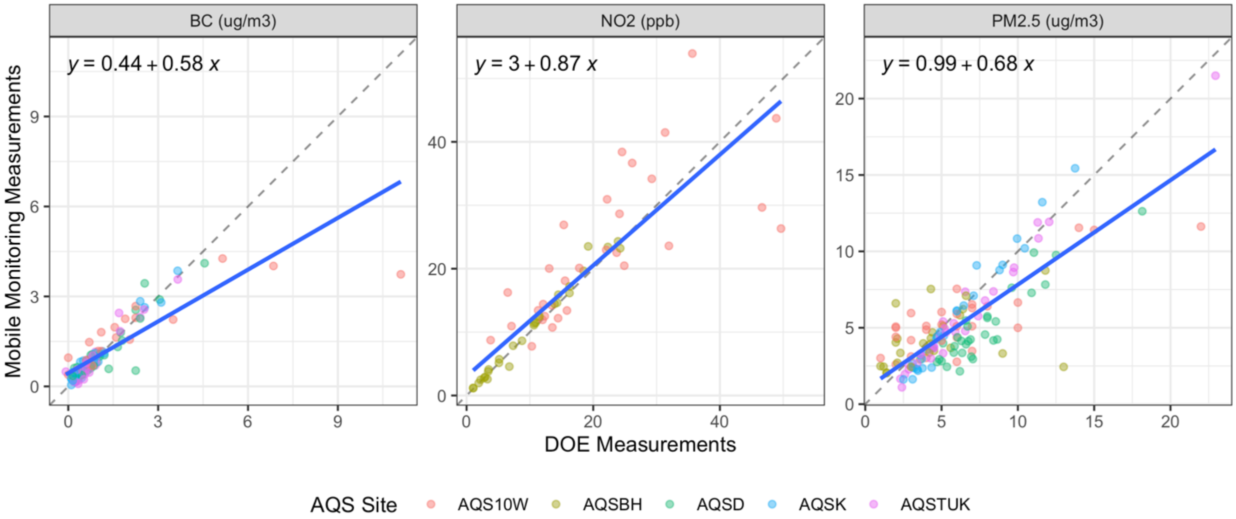

*Figure S12. Comparison of two-minute median concentrations from mobile monitoring and the Department of Ecology (DOE) readings at air quality system (AQS) collocation sites. MSE-based $R^2$: BC = 0.69, $NO_2$ = 0.71, $PM_{2.5}$ = 0.61. The dashed line is the 1-1 line; the blue line is the least squares linear regression fit. Mobile monitoring $PM_{2.5}$ concentrations are from calibrated nephelometer readings (see Methods). DOE $PM_{2.5}$ concentrations are from nephelometers when available (AQSD, AQSK, AQSTUK – readings are updated every minute), otherwise they are from gravimetric and beta attenuation (BAM) methods, which are updated less frequently (AQS10W – readings are based on rolling 1-hour estimates updated every 6 minutes, AQSBH – readings are updated hourly).*

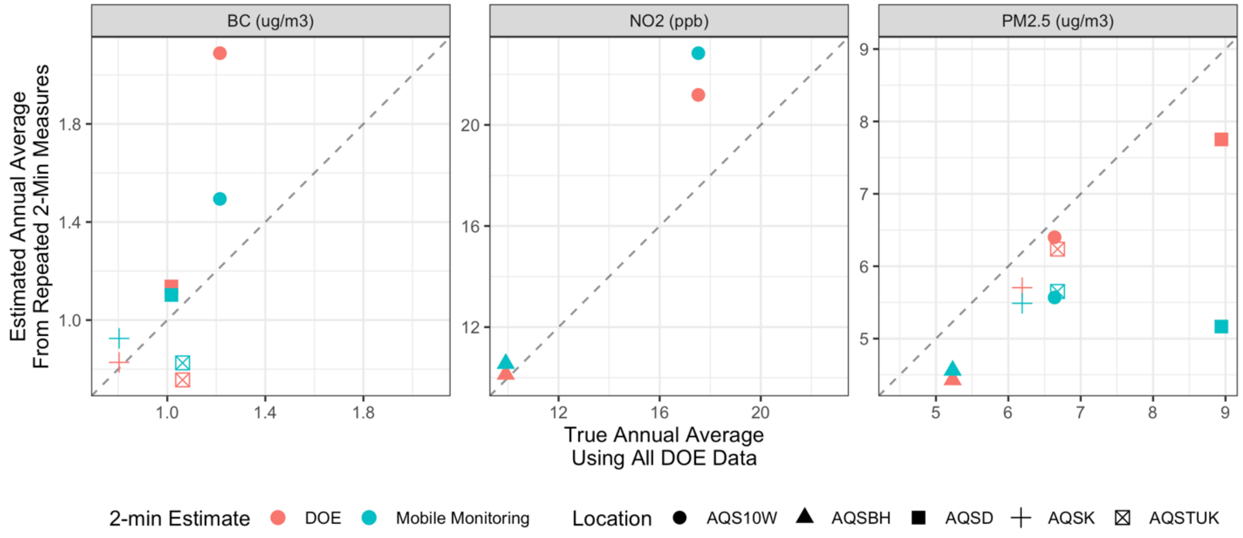

*Figure S13. Comparison of true annual average pollutant estimates at air quality system (AQS) collocation sites to annual average estimates from repeated 2-min measures from mobile monitoring and the Department of Ecology (DOE). Plots compare estimates using mobile monitoring stop data, DOE data during the same two-minute time periods, to the true annual averages at those sites using all the available regulatory monitoring data for the study period.*

#### S2.3 Spatial and Temporal Variability

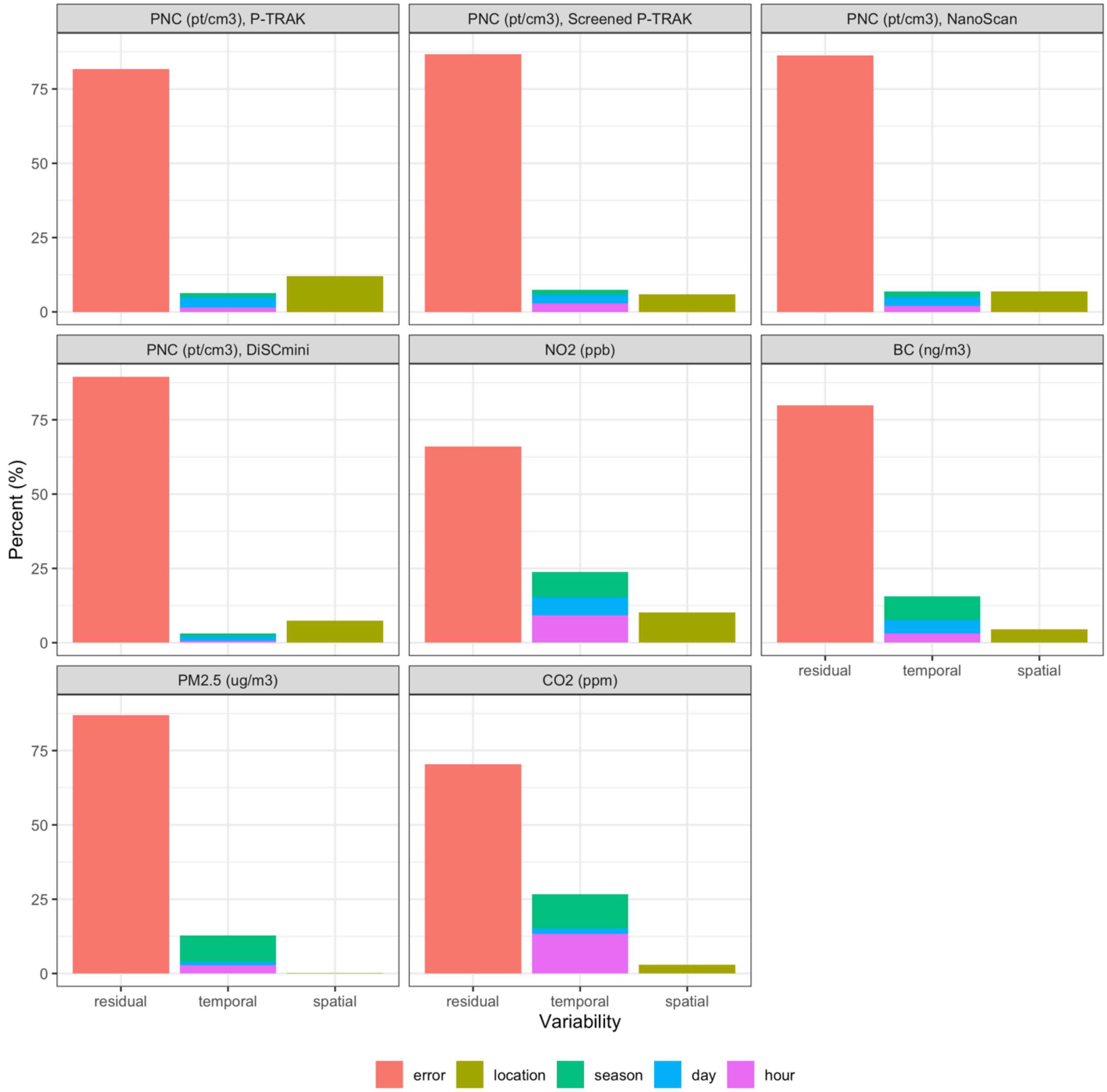

*Figure S14. Percent of winsorized median visit concentration variability explained by spatial, temporal and within site factors for each pollutant (total = 100%). Pollutant values are based on separate Analysis of Variance (ANOVA) models. Spatial variability is the concentration variability across 309 sites. The residual error term represents within-site variability across approximately 29 visits per site.*

#### S2.4 Annual Averages

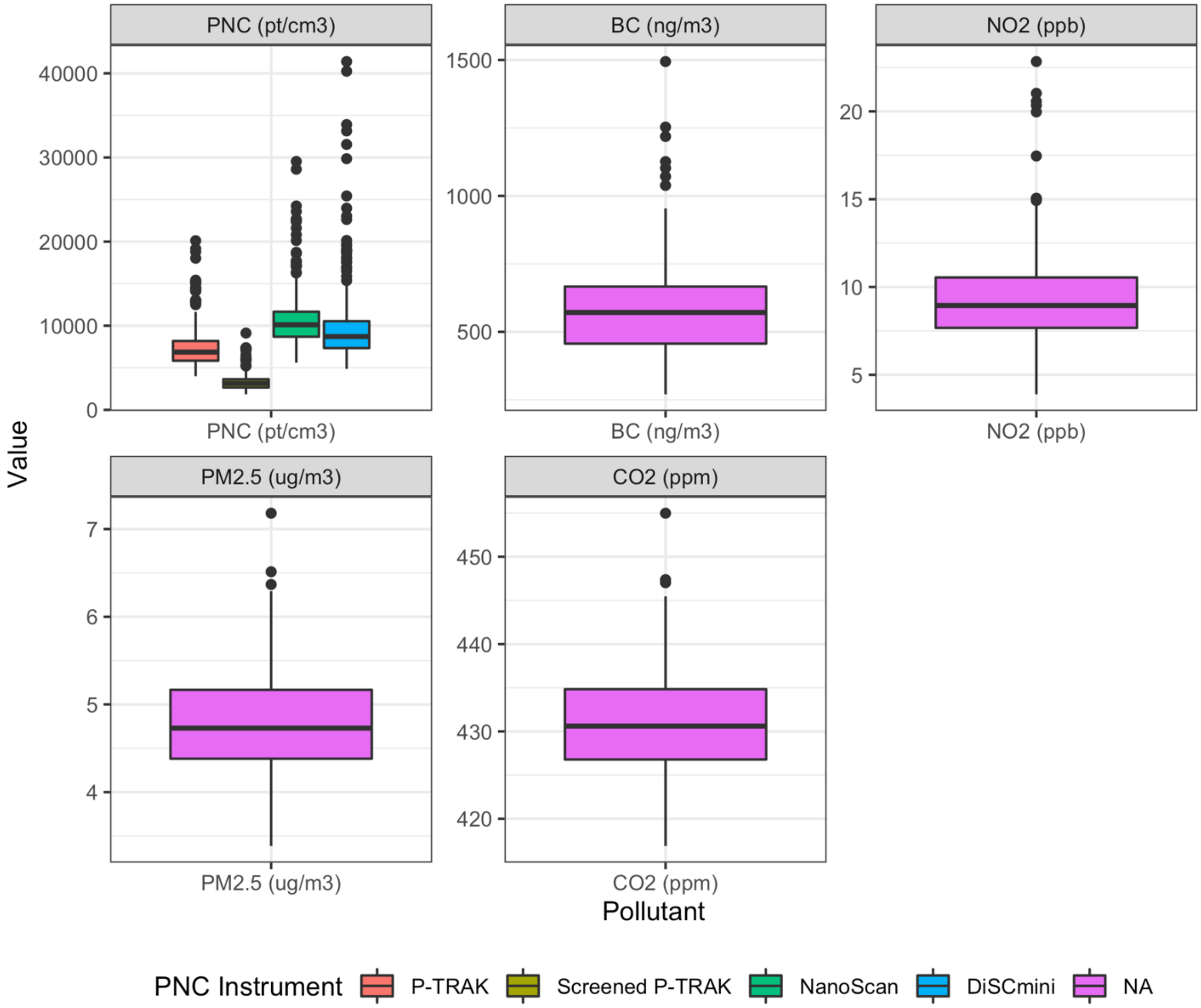

*Figure S15. Annual average site concentrations from winsorized median visit concentration (N=309). The "NA" PNC legend value refers to pollutants other than PNC.*

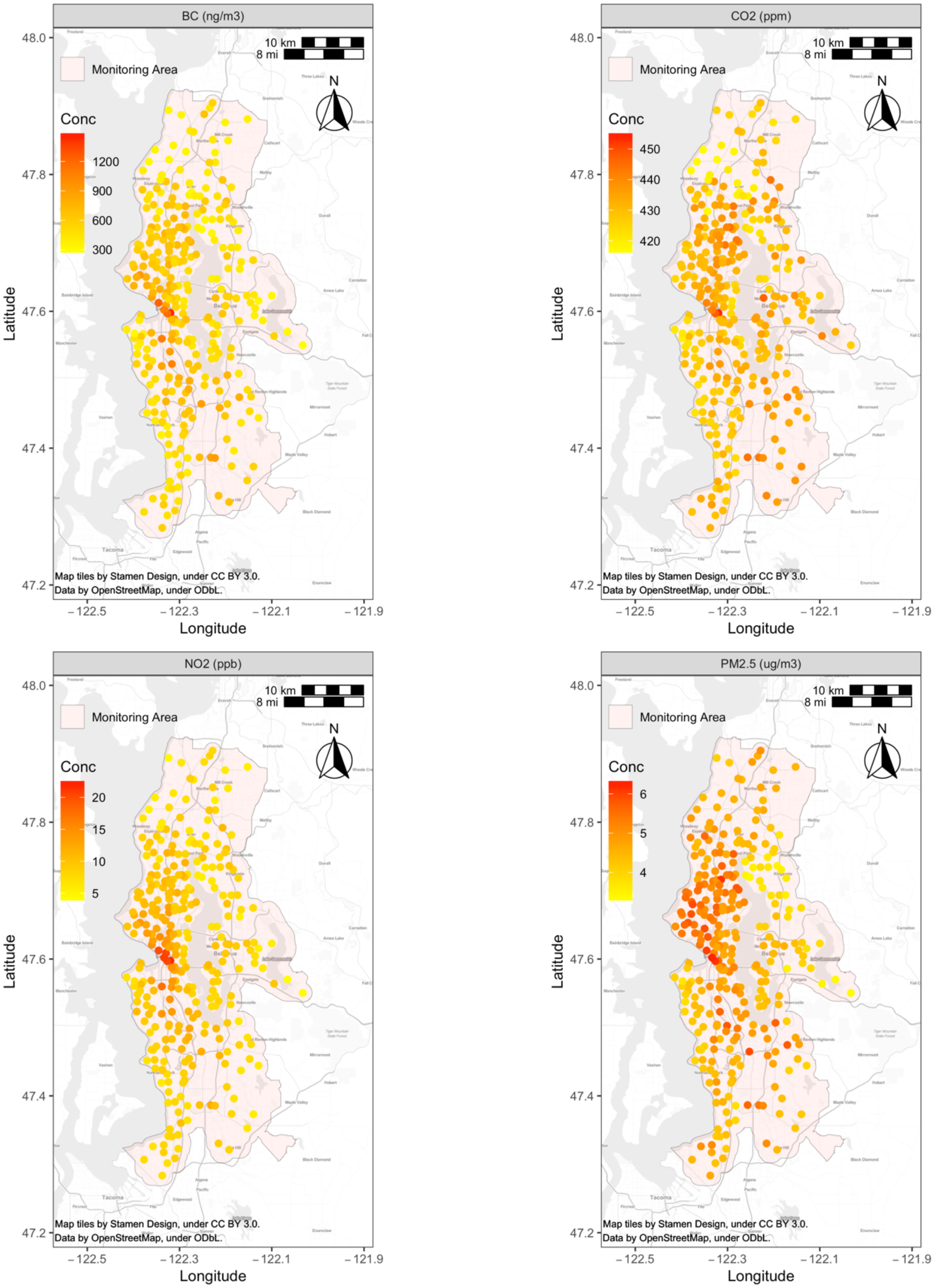

*Figure S16. Annual average $PM_{2.5}$, BC, $NO_2$ and $CO_2$ concentrations at monitoring sites (N=309).*

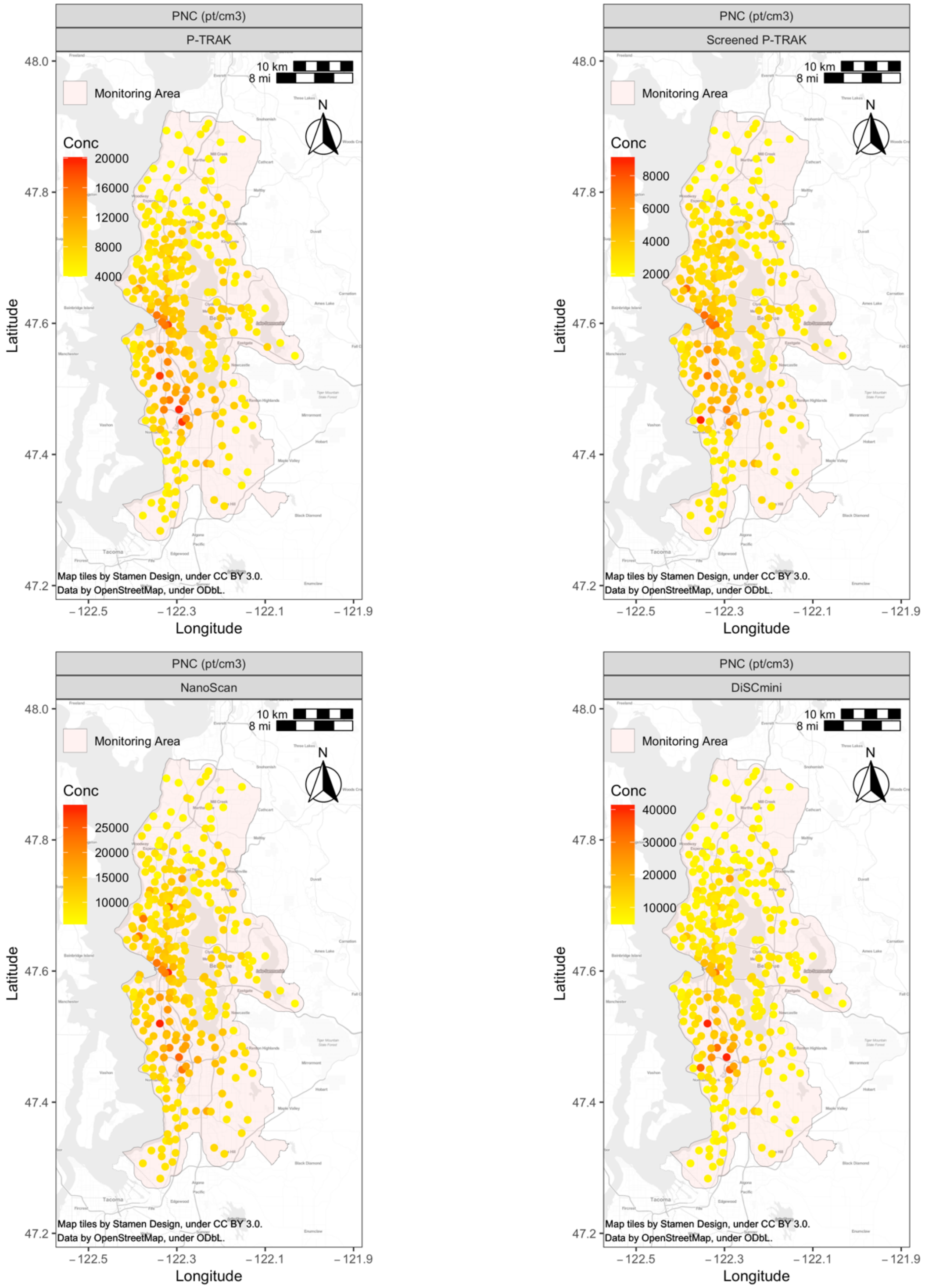

*Figure S17. Annual average PNC concentrations at monitoring sites (N=309) from different PNC instruments.*

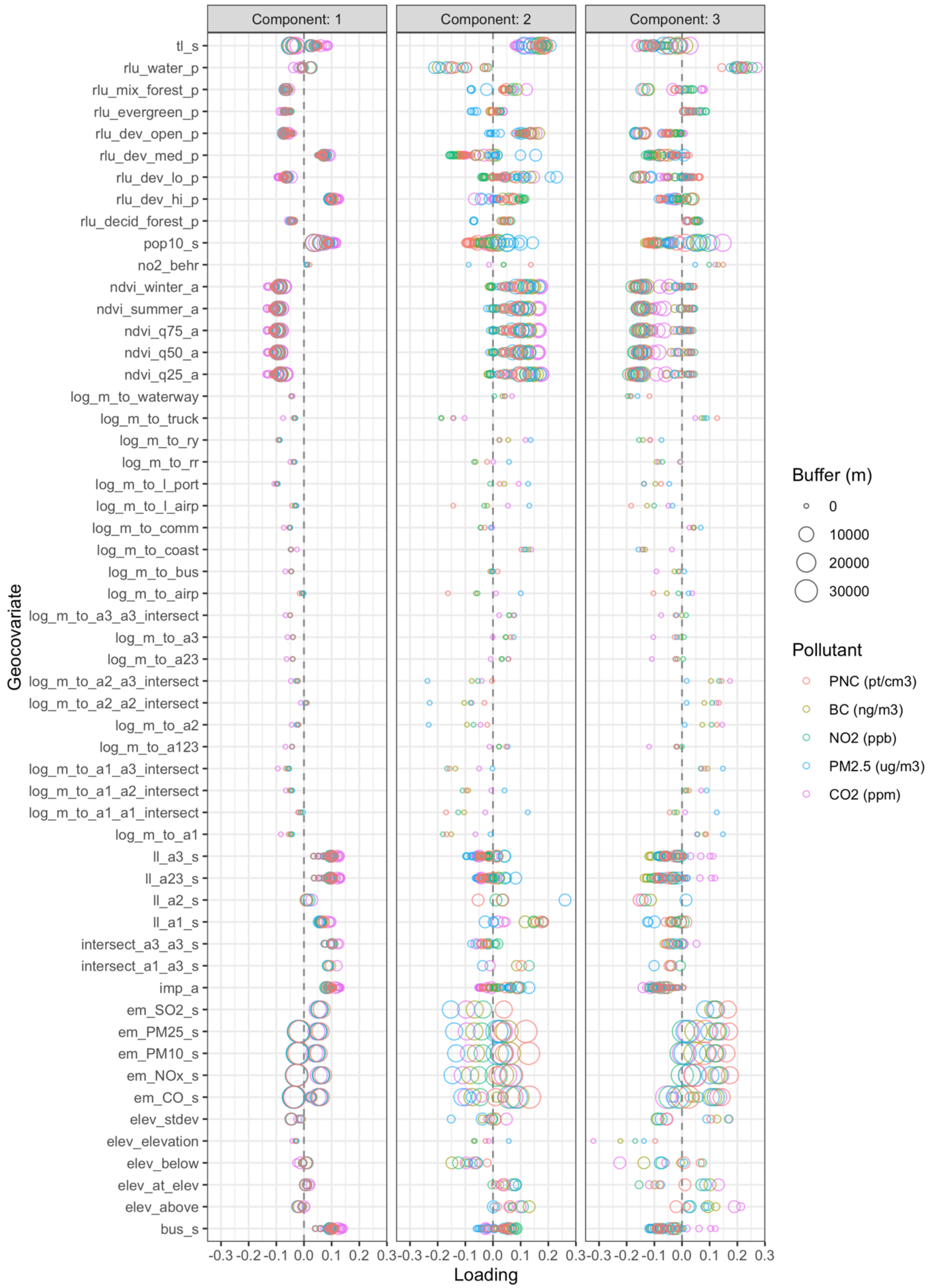

*Figure S18. PLS loadings for pollutant models. PNC results are from the primary instrument, the P-TRAK.*

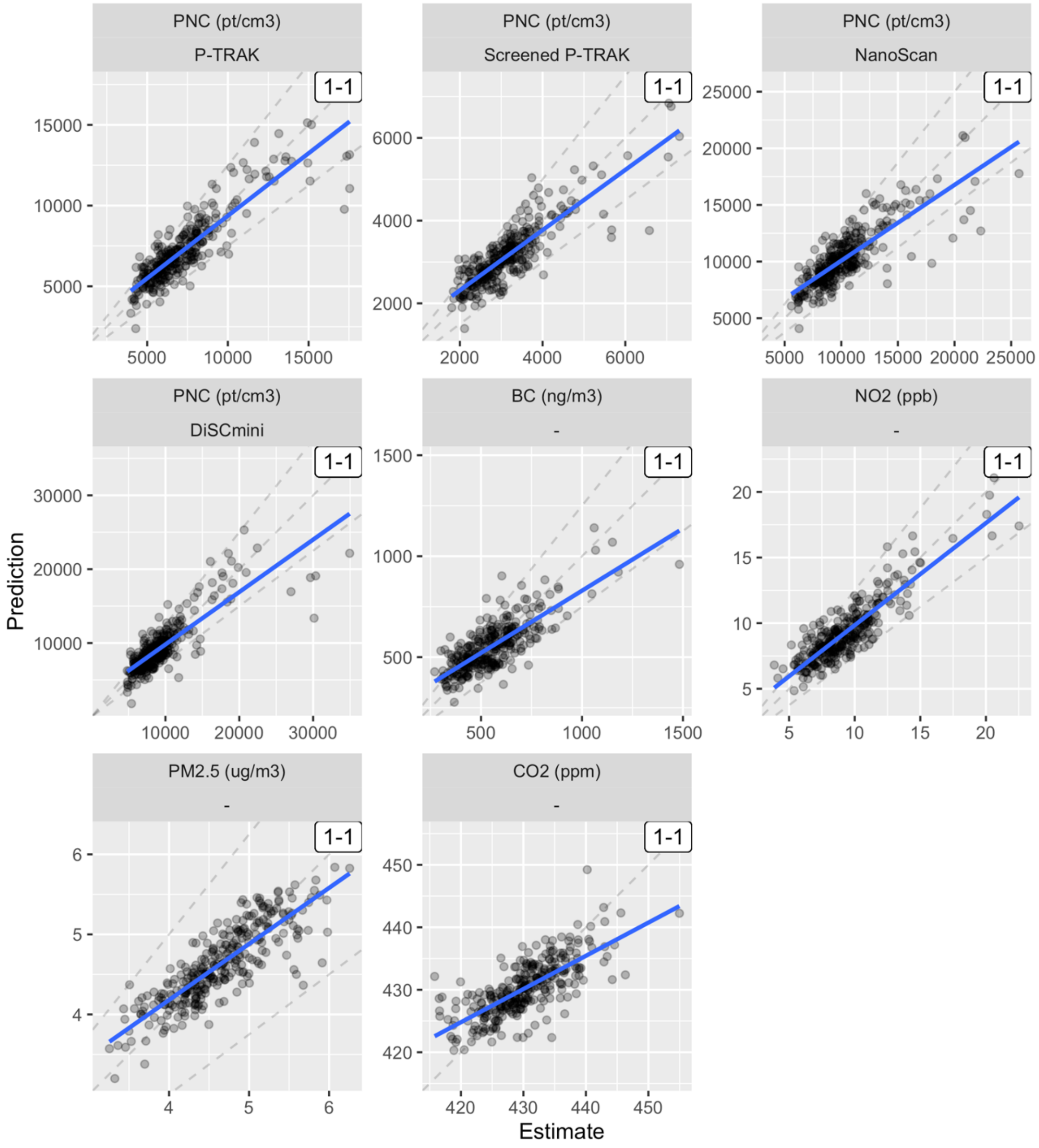

*Figure S19. UK-PLS model predictions of annual average pollutant concentrations. Dashed lines indicate the 1-1 line, as well as 25% above and below (note that $CO_2$ has a narrow range). The blue line shows the best fit line.*

*Table S9. Out-of-sample (OOS) model performances for annual average prediction models at cross-validation (CV; N=278) and test (N=31) sites. The mean of winsorized medians is the primary analysis.*

| Pollutant | | OOS | MSE-based R2 | | | RMSE | | |
|---|---|---|---|---|---|---|---|---|
| | | | **Mean of Winsorized Medians** | **Mean of Medians** | **Median of Medians** | **Mean of Winsorized Medians** | **Mean of Medians** | **Median of Medians** |
| PNC ($pt/cm^3$) | P-TRAK | CV | 0.77 | 0.74 | 0.79 | 1177 | 1320 | 884 |
| PNC ($pt/cm^3$) | P-TRAK | Test | 0.78 | 0.75 | 0.76 | 815 | 882 | 810 |
| PNC ($pt/cm^3$) | Screened P-TRAK | CV | 0.72 | 0.48 | 0.71 | 473 | 738 | 390 |
| PNC ($pt/cm^3$) | Screened P-TRAK | Test | 0.80 | 0.74 | 0.82 | 303 | 363 | 253 |
| PNC ($pt/cm^3$) | NanoScan | CV | 0.65 | 0.47 | 0.69 | 1819 | 2596 | 1404 |
| PNC ($pt/cm^3$) | NanoScan | Test | 0.75 | 0.61 | 0.74 | 1027 | 1327 | 994 |
| PNC ($pt/cm^3$) | DiSCmini | CV | 0.69 | 0.44 | 0.74 | 2339 | 3807 | 1304 |
| PNC ($pt/cm^3$) | DiSCmini | Test | 0.63 | 0.42 | 0.74 | 1390 | 1788 | 1100 |
| BC ($ng/m^3$) | - | CV | 0.60 | 0.60 | 0.58 | 102 | 110 | 78 |
| BC ($ng/m^3$) | - | Test | 0.80 | 0.61 | 0.59 | 60 | 94 | 60 |
| $NO_2$ (ppb) | - | CV | 0.77 | 0.77 | 0.72 | 1.3 | 1.3 | 1.4 |
| $NO_2$ (ppb) | - | Test | 0.84 | 0.85 | 0.72 | 0.9 | 0.8 | 1.1 |
| $PM_{2.5}$ ($\mu g/m^3$) | - | CV | 0.70 | 0.60 | 0.62 | 0.3 | 0.4 | 0.3 |

| | | | | | | | | |
|---|---|---|---|---|---|---|---|---|
| $PM_{2.5}$ (µg/m$^3$) | - | Test | 0.73 | 0.58 | 0.71 | 0.3 | 0.4 | 0.2 |
| $CO_2$ (ppm) | - | CV | 0.51 | 0.51 | 0.37 | 4.2 | 4.2 | 4.4 |
| $CO_2$ (ppm) | - | Test | 0.77 | 0.77 | 0.38 | 2.7 | 2.8 | 4.3 |

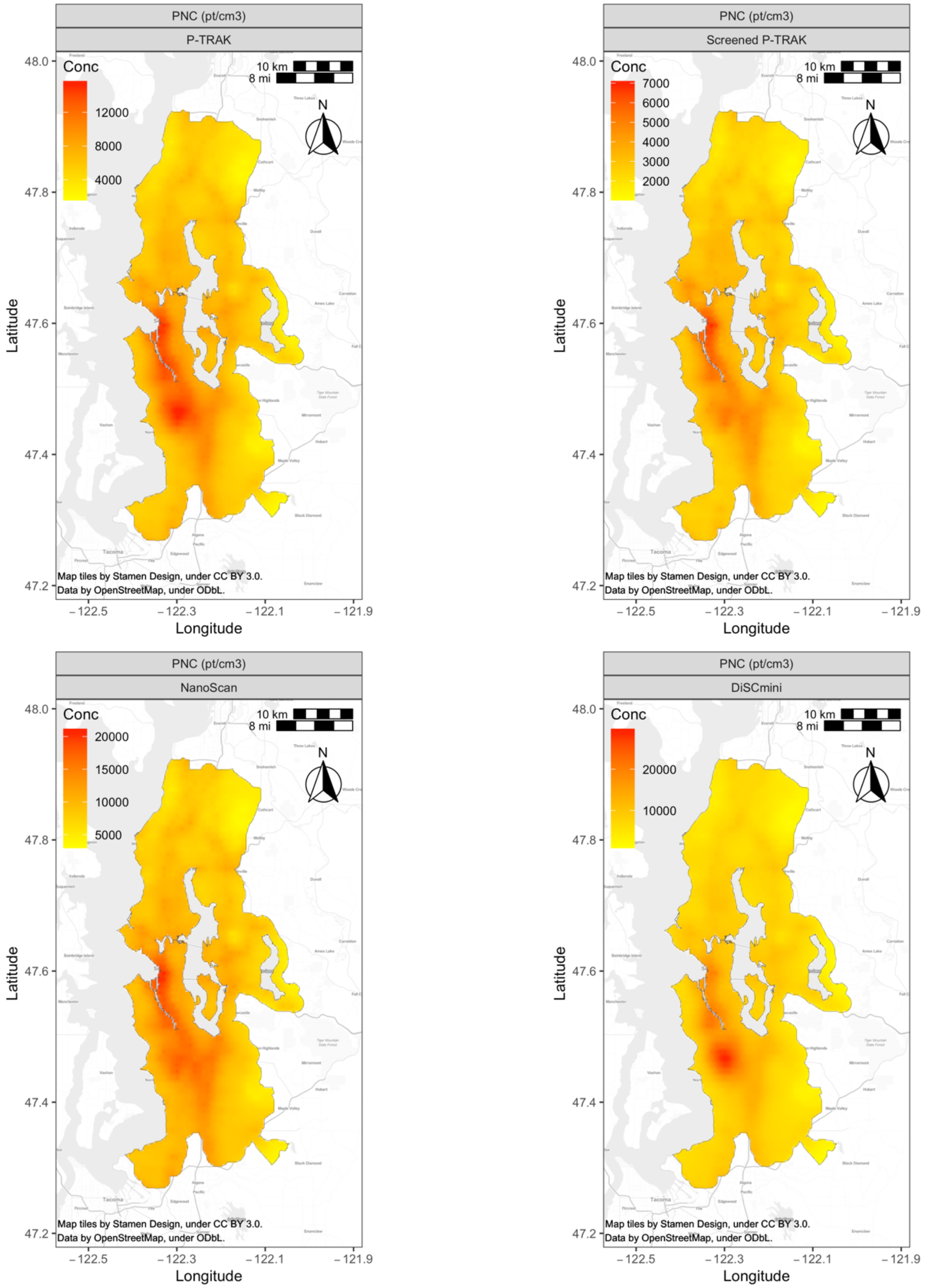

*Figure S20. UK-PLS pollutant predictions within the monitoring region for all PNC instruments. The P-TRAK is the primary PNC instrument.*

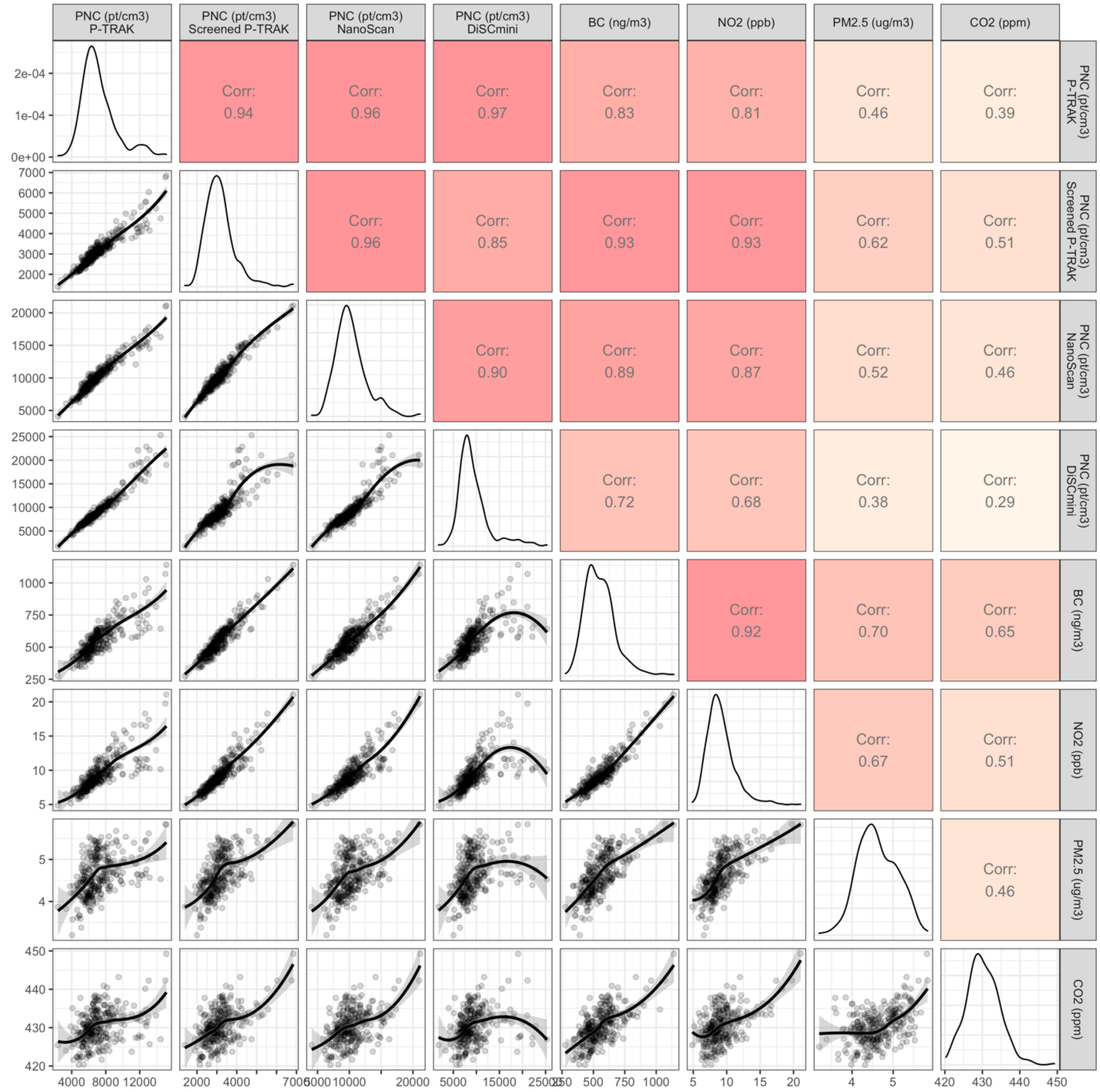

*Figure S21. Annual average pollutant prediction correlations (N=309 sites). Lower panels show scatterplots with loess lines and 95% confidence intervals; upper panels show Pearson correlations (R), with higher values in darker reds; diagonal panels show density plots.*

### S3 Discussion

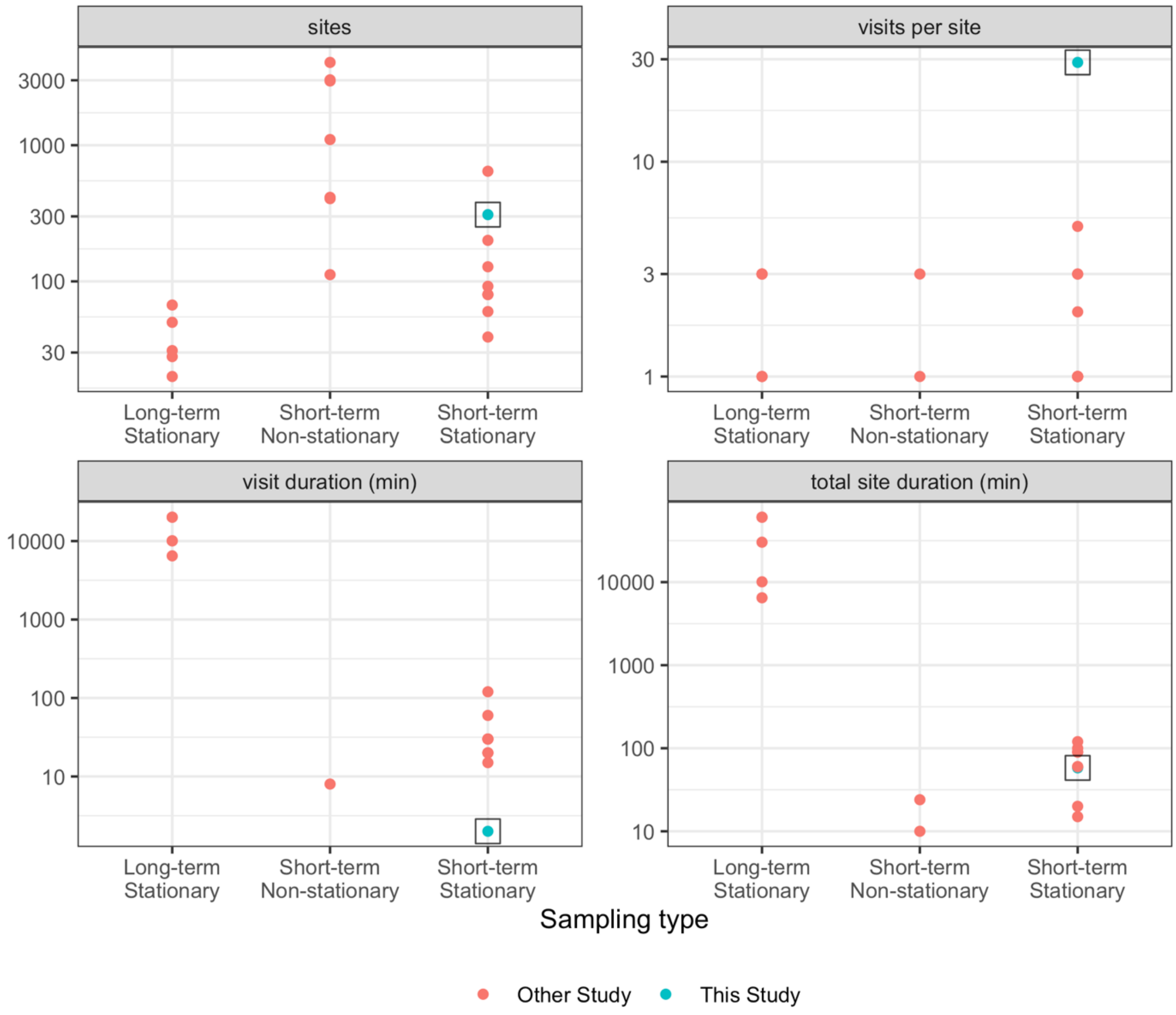

*Figure S22. Sampling approaches across our and other PNC studies.[46–69] Studies are stratified by whether the sampling type was traditional, fixed site sampling (long-term stationary), short-term mobile monitoring campaigns that collected on-road data while in motion (short-term non-stationary), or short-term mobile monitoring campaigns that collected data while stopped (short-term stationary). Figure does not include Saha et al. (2021), who used a mixed sampling approach for PNC from multiple sources.[70] Note that little data were available for short-term non-stationary studies regarding visit duration, total site duration or visits per site. The single study under short-term non-stationary visit duration of ~ 8 min was conducted with pedestrians (Sabaliauskas et al. 2015).*

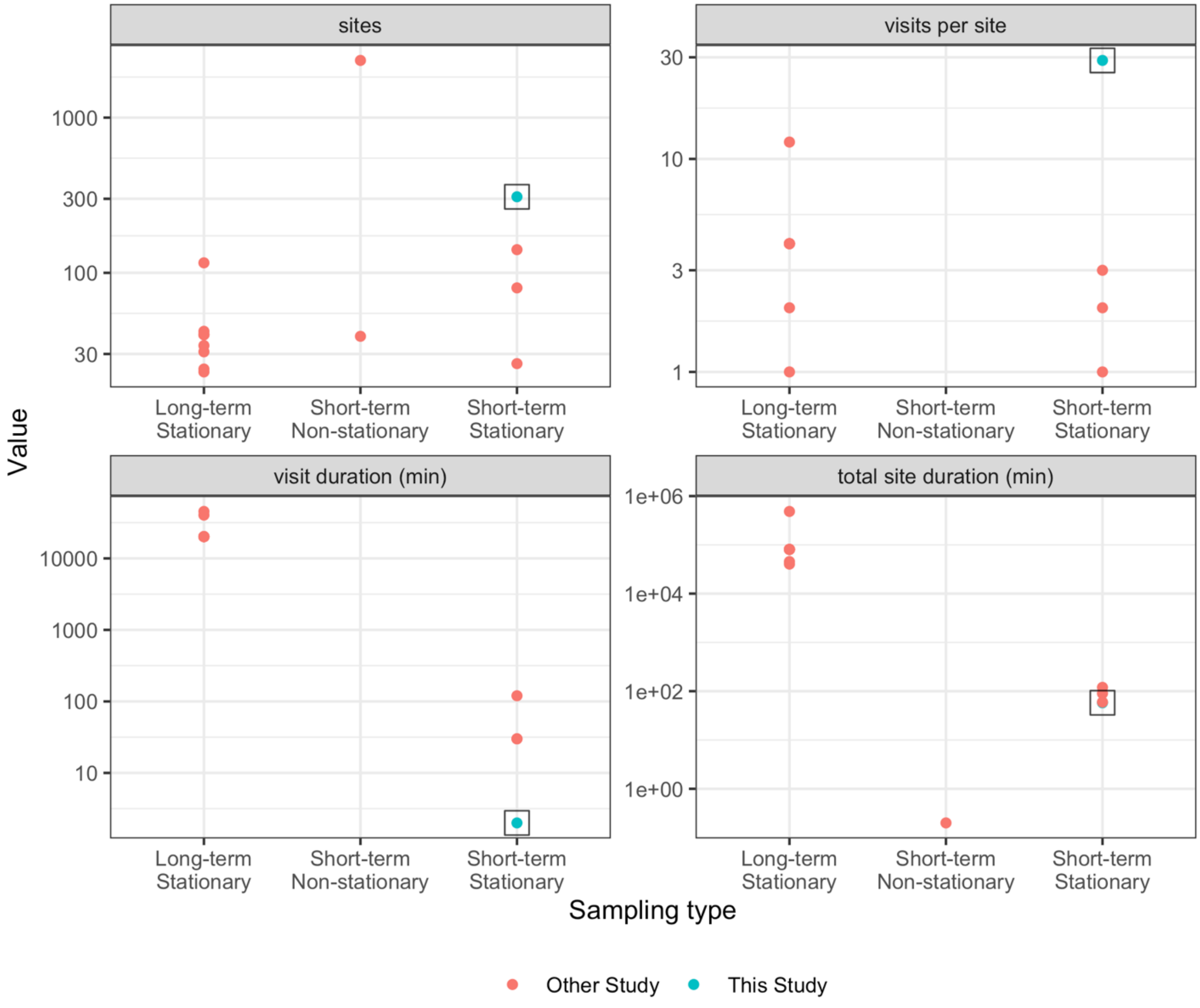

*Figure S23 Sampling approaches across other BC studies.*[46,51,56,61,62,71–81] *Studies are stratified by whether the sampling type was traditional, fixed site sampling (long-term stationary), short-term mobile monitoring campaigns that collected on-road data while in motion (short-term non-stationary), or short-term mobile monitoring campaigns that collected data while stopped (short-term stationary). Note that little data were available for short-term non-stationary studies regarding visit duration, total site duration or visits per site.*

45. Keller JP, Olives C, Kim SY, et al. A unified spatiotemporal modeling approach for predicting concentrations of multiple air pollutants in the multi-ethnic study of

atherosclerosis and air pollution. *Environ Health Perspect*. 2015;123(4):301-309. doi:10.1289/ehp.1408145
